# Effects of an infant formula containing a whey protein concentrate on feeding tolerance and markers of intestinal immune defense in Chinese infants

**DOI:** 10.64898/2026.02.11.26345996

**Authors:** Ying Wang, Min Liu, Shaillay Kumar Dogra, Karine Vidal, Jean-Philippe Godin, Noura Darwish, Xiaona Wei, Lucas Reymond, Qiaoji Li, Jie Dong, Aikaterini Themis Vyllioti, Jodi Bettler, Elaine Kennedy, Keqing Wang, Qiangrong Zhai, Jonathan O’Regan, Tinu Mary Samuel, Wei Cai

## Abstract

**Background:** Human milk (HM) bioactive components can have immune modulatory functions, impact the gut microbiome, and may result in functional benefits when added to infant formula (IF). In this single-arm, prospective, intervention study, we tested the effectiveness of an IF with a whey protein concentrate co-enriched in α-lactalbumin, milk fat globule membrane (MFGM), and *Sn-2* palmitate resulting in protein and lipid profiles observed in HM. The outcomes tested were feeding tolerance, *Bifidobacteria* abundance, and intestinal and immune health of Chinese infants.

**Methods:** Predominantly formula-fed (FF) and breastfed (BF) infants were enrolled between 3 and 28 days and assigned to the FF (N= 60) or BF (N=60) group, per their feeding practice, for 6 weeks. The primary endpoint was Infant Gastrointestinal Symptom Questionnaire (IGSQ) index score assessed using a validated IGSQ-13 questionnaire after 6 weeks of intervention; non-inferiority of FF vs BF was tested. Secondary endpoints included fecal *Bifidobacteria* abundance assessed using shotgun metagenomics sequencing; fecal short chain fatty acids (SCFAs) analyzed by ultra-performance liquid chromatography-tandem mass spectrometry; fecal markers of immune response, inflammation, intestinal barrier integrity (secretory immunoglobulin A sIgA), cytokines, calprotectin, α1 antitrypsin, lipocalin-2) assessed using enzyme-linked immunosorbent assay; stool consistency assessed using gastrointestinal (GI) diary; anthropometric assessments; quality of life; physician reported adverse events; and use of medications.

**Results:** Good GI tolerance was observed in both groups at V2 (mean±SD IGSQ score FF: 19.9±7.4; BF: 16.8±4.2); difference of means 1.35 [95% CI: -1.312, 4.012]). After 6 weeks, *Bifidobacterium* genus relative abundance was not significantly different between the groups. Total SCFAs were significantly higher (p<0.05) in the FF versus BF group, driven by increased levels of valeric and propanoic acids (p<0.05 for both). The IGSQ domain scores, stool consistency, fecal markers of immunity, inflammation, and intestinal barrier integrity (except lipocalin-2 which was significantly higher in BF vs FF), anthropometric Z-scores, common illnesses, antibiotic use, and adverse events were not significantly different between groups at week 6.

**Conclusions:** Our results support the effectiveness of this tested infant formula in supporting good GI tolerance, growth, specific intestinal and immune health markers, and *Bifidobacteria* abundance similar to that of the BF group.

**Trial registration:** NCT04880083 (2021-05-06)

## Introduction

Human milk (HM) is recognized as the normative standard for infant feeding (1). The nutritional components of HM can be categorized into (i) macronutrients (e.g., proteins, lipids, and carbohydrates including oligosaccharides) and (ii) micronutrients (e.g., vitamins and minerals) (2, 3). In addition, HM also contains distinct bioactive molecules that are shown to protect against infection and contribute to immune and gut development (2, 3). Collectively, these components support normal growth and development as well as provide important metabolic, immune, and other biological benefits to the infant (2, 3).

The total protein content of HM is ∼10g/L and it is estimated to contain over 400 different proteins (4); the protein profile of HM can be broadly categorized into three groups: whey proteins, caseins, and milk fat globule membrane (MFGM) proteins (3), the latter contributing to only 1–4% of the total protein (5). These proteins play a critical role in physiological and biochemical mechanisms and in turn in the development and health of an infant.

The whey protein fraction of HM is dominated by α-lactalbumin, lactoferrin, lysozyme, sIgA, serum albumin, and osteopontin. In addition to providing essential amino acids and peptides for growth and development, these proteins also confer other health benefits. For example, the major whey protein α-lactalbumin (which constitutes about 25% of the total protein and 36% of the total whey protein in HM) has been shown to impact gastric emptying time, mineral absorption and delivery, and inhibition, of potential pathogens, *in vitro* and *in vivo* (3). Although HM contains a much lower level of the protein osteopontin compared to standard bovine milk-based infant formulas, it plays a role in infant intestinal, immune, and brain development which demonstrates the biological importance of this protein fraction (6).

The casein protein fraction of HM is dominated by β-casein (A2 variant) and κ-casein with lower levels of α-casein compared to bovine milk (7). These casein proteins are present as casein micelles and play a role in curd formation in the infant stomach as well as being an essential carrier of minerals such as calcium and phosphate. HM does not contain β-lactoglobulin or αS_2_-casein, but has much higher concentrations of β-casein, α-lactalbumin, osteopontin, and lactoferrin than bovine milk. For this reason, creating novel protein fractions (typically from bovine milk) that simulate the composition, structure, and function of HM proteins has been a major challenge for infant formula (IF) innovation.

Lipids are the predominant source of energy in HM, providing close to 50% of the energy intake of breastfed (BF) infants in the first 6 months of life (8). Aside from being the main source of energy for general growth of the infant, lipids also serve specific roles which contribute to neurodevelopment, gastrointestinal (GI) function, and immunity (9). The lipids in HM are present in the form of globules, predominantly (∼98% of total lipids) composed of triacylglycerol (TAG), with the remaining 2% as monoglycerides, diglycerides, sterols, and non-esterified fatty acids (10). These lipid globules are dispersed throughout the milk and stabilized by a complex tri-layered membrane known as the MFGM. The TAG in HM contains a range of fatty acids with palmitic, oleic, and linoleic acids being the dominant components and in a distinctive structure. In HM, approximately 70% of the palmitic acid is esterified to the *Sn-2* position of the triacylglycerol molecule (11). Lipid blends, enriched in palmitic acid that are esterified in their *Sn-2* position, are used in IF. A study showed that when a lipid blend in the IF was enriched in *Sn-2* palmitate, it led to reduced stool soaps, softer stools, and increased *Bifidobacteria*. Adding oligofructose (OF), to the same formula matrix further improved stool consistency resulting in fewer formed stools versus a control formula and brought the GI profile of the formula fed infants closer to that of BF infants (12).

The tri-layered membrane of MFGM contains many lipids (including polar lipids and glycolipids) and proteins (including bioactive glycoproteins and indigenous enzymes). Amongst the lipids of the MFGM membrane are the phospholipids (PLs: phosphatidylcholine [PC], phosphatidylethanolamine [PE], phosphatidylserine [PS], and phosphatidylinositol [PI]), and the major sphingolipids which include sphingomyelin (SM). These PLs are present in all cell membranes and play a wide variety of structural and biological functions including lipid digestion, absorption, and transport (13, 14), as well as cellular processes such as myelination of the central nervous system (15, 16). Cerebrosides and gangliosides are also key components of the MFGM. Gangliosides are thought to positively impact GI and immune health by two main mechanisms: influencing the gut microbiota to promote the growth of *Bifidobacteria*, and by binding to pathogenic targets in the intestine to reduce infections (17, 18). In addition to these bioactive components, HM MFGM also contains over 300 proteins (19). The major proteins of the MFGM include butyrophilin, xanthine dehydrogenase/oxidase, adipophilin, periodic acid Schiff 6/7, mucin 1, and fatty acid binding protein. These proteins have been shown to exhibit a variety of biological functions including the promotion of immune and intestinal development and maturation (20).

The advancement of process technologies has enabled generating dairy ingredients that are enriched in protein fractions such as α-lactalbumin, lactoferrin, osteopontin, and MFGM (20–26), allowing the design of new formulas to confer improved functional benefits to formula-fed infants. Development of such process technologies is a key contributor to enabling IF to be as close as possible to HM. In a randomized clinical trial (RCT) with a standard IF, an experimental formula enriched with α-lactalbumin, and non-randomized BF group, the GI tolerance profile of α-lactalbumin-enriched formula-fed infants was similar to the breastfed group (23). In another study, when healthy term infants were randomized to three IF groups with varying levels of osteopontin (0, 65, or 130 mg/L) and compared to a non-randomized BF group, lower levels of the systemic proinflammatory cytokine tumour necrosis factor-alpha (TNF-α) at ages 1 and 4 months and a lower frequency of fever were reported in the osteopontin-supplemented formula groups (27). Separately, in a multi-center RCT, healthy term infants fed a MFGM-enriched formula exhibited better GI tolerance compared to a standard formula group, and growth rates in the first year of life were similar in both formula groups compared to a reference BF group (28).

While these observations suggest that IF enriched with these individual bioactive components may provide health benefits resembling the ones observed in BF groups, additional studies are warranted to document and monitor the real-world effectiveness of consuming IF with a combination of these bioactives on infant health and development for cases when breastfeeding or providing HM is not an option.

The current study evaluated an IF designed with a specific combination of process technologies that result in similar protein and lipid profiles to HM, specifically, an advanced whey protein profile obtained using a whey protein ingredient that delivers a formula enriched in α-lactalbumin, osteopontin, lactoferrin and MFGM proteins and lipids (α-MFGM) (29) and a casein profile comprised of A2 β-Casein, in combination with an *Sn-2* palmitate enriched TAG core. This was a single-arm, open-label, prospective interventional study in a real-world setting to examine the effectiveness of the formula in promoting good GI tolerance and intestinal and immune health outcomes in healthy term Chinese infants compared to a BF reference group.

## Methods

### Study design and population

This multi-center, single-arm, open-label, prospective intervention study was conducted between June 2021 and October 2022 in Xinhua Hospital, School of Medicine, Shanghai Jiao Tong University and Shanghai Public Health Clinical Center in China. A total of 122 neonates were screened, where legal guardians provided written informed consent before assessment against all protocol specified inclusion and exclusion criteria. Two neonates (1.6%) were screen failures as their parent(s) or legal representative were not willing and able to comply with scheduled visits and the requirements of the study protocol. All eligible neonates were then scheduled for the baseline visit, where subject numbers were assigned, screening records completed, anthropometric measurements obtained, questionnaires (IGSQ 13 and IQI) administered, fecal sample kits and feeding logs provided, and follow up visits scheduled. Enrolment was confirmed once all baseline procedures and data collection were completed. Infants were enrolled between 3 and 28 days of age if they met the following criteria: healthy, term infant (37-42 weeks of gestation); birth weight ≥ 2500g and ≤ 4500g; FF infants were to predominantly consume and tolerate a standard cow’s milk IF, and their parents must have independently elected to not exclusively breastfeed before enrollment; BF infants must have been predominantly breastfed since birth or since ≥ 7 days (in case they have switched from FF to BF), and their parents must have elected predominant breastfeeding for the study duration. Infants were considered as predominantly breastfed when fed with breast milk for at least 75% of total milk feeds per day. Exclusion criteria were as follows: infants receiving complementary foods or liquids defined as 4 or more teaspoons (∼20g) per day; those with conditions requiring other infant feedings; infants with evidence of congenital malformations, documented systemic or congenital infections, or with previous or ongoing severe medical or laboratory abnormalities; those receiving or have received probiotic supplements or medications/supplements known or suspected to affect fat digestion, absorption, metabolism, stool characteristics and microbiota, growth, and gastric acid secretion; and infants currently participating or having participated in another clinical trial since birth.

This study was approved by the ethics committees at the two institutions in China, which were the Ethics Committee of Xinhua Hospital Affiliated to Shanghai Jiao Tong University School of Medicine (XHEC-C-2020-053-1), and the Ethics Committee of Shanghai Public Health Clinical Center (2020-E151-01), respectively and conducted in compliance with the Declaration of Helsinki and the International Conference on Harmonization Guidelines for Good Clinical Practice. Prior to enrollment, parent(s)/legally authorized representative provided written informed consent. This trial was registered on ClinicalTrials.gov (NCT04880083) and carried out according to the Consolidated Standards of Reporting Trials (CONSORT) guidelines. The completed CONSORT checklist is provided in ***Additional file 1*.**

### Intervention

In this single-arm, open label, prospective interventional study, FF infants were compared to a parallel-recruited BF infant group. The FF infants began the intervention one day after the baseline visit (V1) and the intervention continued for 6 weeks until the study end (V2). The BF infants continued with their breastfeeding regimen until V2.

#### Study Product

The study formula was a bovine milk-based IF manufactured by Wyeth Nutritionals Ireland, Askeaton, Ireland and commercially sourced in China. The whey protein dominant IF contained a specific whey protein profile that delivers a formula enriched in α-lactalbumin, osteopontin, lactoferrin and MFGM proteins and lipids and a casein profile of A2 β-casein, in combination with an *Sn-2* palmitate enriched TAG core (**Table 1**). The formula was provided in cans in powder form with a commercial label and tag for clinical trial use only. Formula was fed orally, ad libitum, via an infant feeding bottle. Intake of formula and breastmilk varied based on the child’s age, weight, and appetite.

**Table 1.**
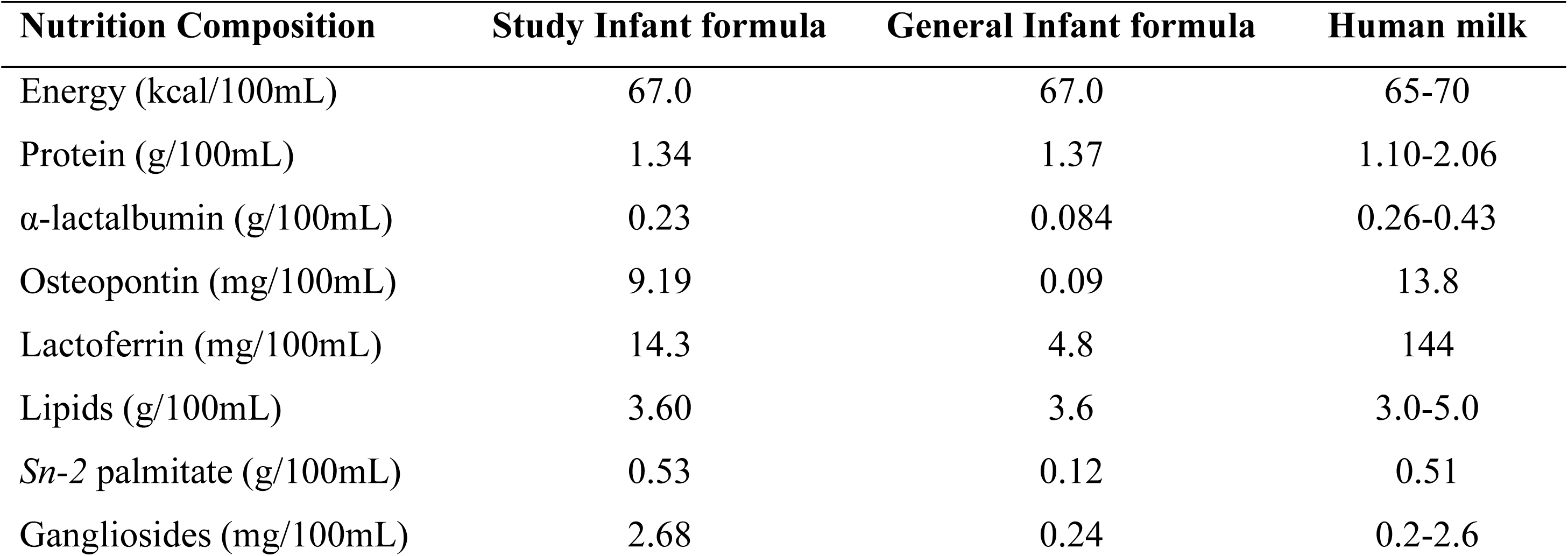
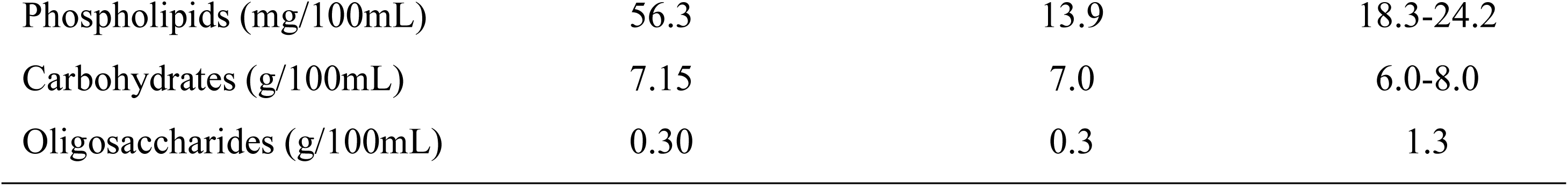
Composition of study formula in comparison to general infant formula (12, 29–32) and human milk (2, 3, 11, 30, 32–34)

| Nutrition Composition | Study Infant formula | General Infant formula | Human milk |
| --- | --- | --- | --- |
| Energy (kcal/100mL) | 67.0 | 67.0 | 65-70 |
| Protein (g/100mL) | 1.34 | 1.37 | 1.10-2.06 |
| α-lactalbumin (g/100mL) | 0.23 | 0.084 | 0.26-0.43 |
| Osteopontin (mg/100mL) | 9.19 | 0.09 | 13.8 |
| Lactoferrin (mg/100mL) | 14.3 | 4.8 | 144 |
| Lipids (g/100mL) | 3.60 | 3.6 | 3.0-5.0 |
| *Sn*-2 palmitate (g/100mL) | 0.53 | 0.12 | 0.51 |
| Gangliosides (mg/100mL) | 2.68 | 0.24 | 0.2-2.6 |
| Phospholipids (mg/100mL) | 56.3 | 13.9 | 18.3-24.2 |
| Carbohydrates (g/100mL) | 7.15 | 7.0 | 6.0-8.0 |
| Oligosaccharides (g/100mL) | 0.30 | 0.3 | 1.3 |

### GI tolerance including GI symptoms and GI-related behaviors

The primary endpoint of this study was the Infant Gastrointestinal Symptom Questionnaire-13 (IGSQ-13) index score for overall GI tolerance calculated after 6 weeks of intervention (V2). The IGSQ-13 is a retrospective, standardized, validated questionnaire of GI symptoms and related behaviors that the infant experienced over the past week from a parent/caregiver perspective (35). Thirteen questions were used to assess the frequency and intensity of infant GI signs and symptoms within 5 domains (stooling, vomiting, flatulence, crying, and fussiness). Composite index scores range from 13-65, with lower scores indicating lower discomfort (35). Good tolerability is a score of 13-23 while a score of more than 23-30 indicates some level of GI distress and a score greater than 30 implies not well tolerated. Individual questions representing each of the 5 domains from the IGSQ-13 were also evaluated separately at V1 and V2 as a secondary assessment.

### Stooling patterns

Stool characteristics were assessed using a one-day retrospective intake diary at baseline and a 3-day intake diary prior to V2. Parents/caregivers were asked to recall infant’s bowel movements over each reporting period. Stool frequency was calculated as the mean number of stools per day. For each bowel movement, parents/caregivers recorded yes/no as to whether it was difficult to pass. The number of stools reported as difficult was divided by the total number of stools to calculate the percentage of difficult bowel movements. Parents/caregivers also judged each stool’s consistency based on 5 possible consistencies using pictures provided in the stooling diary. Each stool category was assigned a score based on the following 5-point scale: 1=watery; 2=runny; 3=mushy/soft; 4=firm; 5=hard (36). The sum of each score was divided by the total numbers of stool scores to calculate mean stool consistency scores.

### Infant quality of life

The Infant Quality of Life Instrument (IQI) is a validated, parent-reported questionnaire for assessing quality of life for infants in the first year of life. It includes 7 items related to breathing, mood, sleeping, feeding, skin, interaction, and stooling and is scored from one to 4 for each item, with increasing scores indicating problematic states (e.g., 1=normal breathing, 2=slight breathing problems, 3=moderate breathing problems, 4=severe breathing problems) (37). In addition, individual IQI scores were weighted by the coefficients reported in publication (37) for the levels of the 7 IQI health items for Chinese primary caregivers, and individual scores were summed to calculate the total IQI score. The total IQI score ranges from 0 to -1.12, with 0 indicating no symptoms and -1.12 indicating the worst symptoms. Parents/caregivers completed the IQI at V1 and V2 via a paper-based form.

### Growth

Standardized anthropometric measurements, which included weight (g), length (cm), head circumference (cm), and corresponding Z-scores, were completed at V1 and V2 to evaluate growth. A calibrated electronic weighting scale was used to weigh infants without a diaper or any clothing at the visits. Weight was recorded twice to the nearest 1g. Measurements were repeated until reproduced within 10g and the two weights within 10g were recorded. Infant length was measured using a standardized length board while the infant was dressed in light underclothing or a diaper without shoes or hair ornaments. Length was recorded twice to the nearest 0.1cm. If the two length measurements were not within 0.5cm, the infant was measured a third time and the two measures in closest agreement were recorded. Infant head circumference was measured twice over the most prominent part on the back of the head and just above the eyebrows and recorded to the nearest 0.1cm using a standard non-elastic plastic-coated measuring tape. If the two head circumference measurements were not within 0.2cm, the infant was measured a third time and the two measures in closest agreement were recorded. Lastly, corresponding Z-scores, including weight-for-age, weight-for-length, length-for-age, body mass index (BMI)-for-age, and head circumference-for-age, were calculated using the World Health Organization (WHO) growth standards (38).

### Adverse events and medication use

Adverse events (AEs) were collected throughout the study period. Reported AEs and serious AEs (SAE) included documentation of type, incidence, severity, seriousness, and relation to feeding. Infant illness and infection outcomes and medication use were part of the standard AE reporting for safety assessment. Symptoms/illness categories of interest included lower respiratory tract infections, upper respiratory infections, acute gastroenteritis/infectious diarrhea, otitis media, and fever. Medication type and duration of use for antibiotics, antipyretics, and anti-reflux medications were also collected for the study period.

### Fecal composition

Fresh fecal samples were collected at V1 and V2 from diapers, immediately stored frozen at -20°C after placing into vials, and transferred to Human Metabolomics Institutes (HMI, Inc, Guangdong, China) for analyses of biomarkers and Beijing Genomics Institute (BGI, Shenzhen, China) for analyses of gut microbiota where the samples were stored at -80°C until analyses.

### Fecal Bifidobacteria abundance

Shotgun metagenomics sequencing was used to quantify *Bifidobacteria* abundance. Magpure Stool DNA KF Kit B (Magen, China) was used to extract deoxyribonucleic acid (DNA) from the fecal samples. Covaris E220 (Covaris, Brighton, UK) was used to yield 300 to 700 base pair (bp) of fragmented DNA and the DNA library was generated with a MGIeasy preparation kit (MGI, China) and loaded into a DNA Immunoprecipitation sequencing (DIP-seq platform, MGI, China) for sequencing to receive 40 million paired-end 100bp reads per sample. SOAPnuke (39) was used for sequencing control and preprocessing of FASTQ files. Metagenomic Phylogenetic Analysis (MetaPhlAn3) (40) was used to profile the composition of microbial communities (bacteria, archaea, and eukaryotes) from metagenomic shotgun sequencing data. Five samples were removed, due to the quality control threshold of low raw data size of <5 GB, details of which are provided in ***Additional file 2***.

### Fecal short chain fatty acids (SCFAs) and lactic acid analysis

Fecal SCFAs (acetate, propionate, butyrate, isobutyrate valerate, isovalerate, caproate, ethylmethylacetate, 4-methlyvalerate, 3-hydroxyisovalerate) and lactate quantification was performed through derivatization with nitrophenylhydrazine in 50% aqueous acetonitrile based on previous report (41) using ultra-performance liquid chromatography-tandem mass spectrometry. The final concentrations of the fecal metabolites were normalized with dry stool weight and expressed as absolute concentrations (µmol/g dry feces) for all the metabolites. SCFAs were taken as the sum of acetate, propionate, butyrate, isobutyrate, valerate, isovalerate, and caproate, while branched chain fatty acids (BCFAs) represent the sum of isobutyrate and isovalerate. Each SCFA and BCFA metabolite was also expressed as relative proportions (%) of total SCFAs and total BCFAs, respectively. SCFA values below the lower limit of quantification (LLOQ) were imputed as LLOQ/2, while values above the upper limit of quantification (ULOQ) were defined as the numerical ULOQ value. Metabolites (such as ethylmethylacetate, 4-methlyvalerate, and 3-hydroxyisovalerate) with more than 60% of missing values below the LLOQ, (***Additional file 3***) were excluded from the analysis (42, 43).

### Fecal markers of immune response, inflammation, and intestinal barrier integrity

Fecal markers of mucosal immune response (sIgA), intestinal inflammation (calprotectin, cytokines, and lipocalin-2), and intestinal barrier integrity (α1 antitrypsin) were assessed at V1 and V2. One gram of freeze-dried feces was re-suspended in 5mL of phosphate-buffered saline, homogenized, and after centrifugation (15,000g, 15 min at 4°C), supernatants were collected for analysis. Commercially available enzyme-linked immunosorbent assay kits were used to quantify fecal sIgA (E-EL-H1275c, Elabscience Biotechnology Co. Ltd, Wuhan, China), calprotectin (ab267628, Abcam, Cambridge, UK), α1 antitrypsin (ab108799, Abcam, Cambridge, UK), lipocalin-2 (ab215541, Abcam, Cambridge, UK), and cytokines which included interleukin-1β (IL-1β), interferon-γ (IFN-γ), and tumor necrosis factor -alpha (TNF-α) (respectively: ml301814, ml002277 and ml002095, Shanghai Enzyme-linked Biotechnology Co. Ltd, Shanghai, China), following the manufacturer’s protocol. Values higher than the ULOQ were replaced by the ULOQ, values lower than LLOQ but higher than limit of detection (LOD) were kept, and values below LOD were imputed as missing (***Additional file 4***). Final concentrations of these proteins were normalized by dry stool mass.

### Sample size

The sample size for the study was calculated for the primary objective. To demonstrate non-inferior IGSQ index score after 6 weeks of intervention in the FF group compared to the BF group, a non-inferiority boundary of 3 IGSQ-score points, which is a 30% effect with respect to transition from good to a problematic score of 23, was set up as it was considered a clinically relevant effect. It was assumed that BF scores by one point better than FF, which would result in effective 2 IGSQ-score points and therefore used in the sample size calculation as actual difference, with a power of 80% and an α-level of 2.5%. A standard deviation of 3.4 IGSQ-score-points in the BF group was assumed for the calculation. It was estimated that 48 infants per group would be needed and assuming 20% attrition, the total number of infants required per group was 60.

### Statistical analysis

The full analysis set (FAS) included all infants enrolled who received at least one study feeding. The per protocol set (PPS) included all infants with no major deviations from the protocol including non-compliance to feeding regimen for at least 80% of the study period. A non-compliant day included non-consumption of study feeding, consumption of non-study feedings (any non-study formula in FF; or probiotic-containing formula or probiotic supplements in BF) or unauthorized complementary foods. Since the study was a non-randomized clinical trial, the safety analysis set (SS) was identical to the FAS. The primary endpoint was analyzed in both the FAS and PPS populations. All secondary endpoints were analyzed in the FAS population (for microbiome analysis some samples were further removed for quality purposes as mentioned above and explained in ***Additional file 2***). All safety analyses were conducted in the SS population.

Descriptive statistics were calculated and tabulated for all categorical and continuous variables by visit and feeding group. The primary endpoint of IGSQ score at 6 weeks was analyzed using analysis of covariance (ANCOVA) with feeding group as the main independent variable and correcting for site, baseline score, delivery mode, sex, and baseline age. Additionally, a robust ANCOVA adjusting for the same covariates was performed as a sensitivity analysis to assess the impact of outliers. Non-inferiority was concluded if the upper bound of the two-sided 95% confidence interval (CI) of the mean difference in IGSQ between FF and BF at 6 weeks was less than 3 score-points. For secondary endpoints, continuous variables were analyzed with an ANCOVA. For the analyses on the absolute concentration of short chain fatty acids, a natural log transformation of the response (and baseline) was applied to achieve approximately normally distributed residuals. SCFA and BCFA metabolites expressed as relative proportions (%) of total SCFAs and total BCFAs did not require such transformation. For fecal secretory IgA, cytokine profile, fecal calprotectin and fecal markers of GI health and maturation, a robust version of the ANCOVA was used. Stool frequency at 6 weeks was analyzed with a Poisson regression with the offset of observed days. All secondary endpoint models had feeding group as main independent variable and adjusted for corresponding site, baseline value, delivery mode, sex, and baseline age. Models where the response was the outcome at baseline were not adjusted for the baseline value.

The overall diversity of the fecal microbial community was quantified by Shannon index. Shannon index was analyzed with an ANCOVA with either feeding group or visit as the main independent variable and adjusting for site, baseline Shannon index (except when Shannon index at baseline was the response or when visit was the main independent variable), delivery mode, sex, and baseline age. Furthermore, the fecal community dissimilarities among the 4 study groups defined by feeding regimen and visit time were displayed using Principal Coordinates Analysis (PCoA) and tested with permutational multivariate analysis of variance (PERMANOVA) with 5000 permutations.

Bacteria relative abundances were analyzed with a zero-inflated negative binomial (ZINB) model, which is a two-component model, a first component that models the strictly positive counts using a negative binomial regression with log link function and a zero-inflated component that models the probability of zero count value using a binomial regression with logit link function (i.e. a logistic regression). The bacteria relative abundances in percentage scale (i.e., 0-100%) were multiplied by 100 (to overcome the fact that most abundances were strictly smaller than 0.05%) and then rounded to obtain integer values. Thus, converted bacteria relative abundances were then used in the ZINB model. The count component of the ZINB model had bacteria relative abundance as the response variable, feeding group as main independent variable, and adjusted for baseline bacteria relative abundance (except when bacteria relative abundance at baseline was the model response), baseline age, delivery mode, sex, and site as covariates. The zero-inflated component of the model adjusted for the same covariates as the count model. Estimates of the mean difference between FF and BF, two-sided 95%-confidence intervals and two-sided p-values associated with those estimates, were derived by combining results from both model components and using asymptotic normal approximation.

The change over time in *Bifidobacterium* genus was compared across the intervention and reference groups by ANCOVA adjusting for baseline *Bifidobacteria* relative abundance, baseline age, site, sex and delivery mode. Feeding group was the main independent variable. Estimate of the mean difference between FF and BF, two-sided 95%-confidence intervals and two-sided p-value were derived.

Associations of gut microbiota with fecal organic acids were assessed using Spearman rank correlation heatmaps with Benjamini-Hochberg false discovery rate correction applied to the set of two-sided p-values for a given bacteria’s association with various fecal organic acids.

Statistical significance for the primary endpoint analysis was tested at the one-sided 2.5% level, while for secondary endpoints, statistical significance was tested at the two-sided 5% level. For secondary endpoints, no adjustment for multiplicity was performed. The microbiota analyses were conducted in R [R Core Team, 2014, version 4.3.2 (2023-10-31 ucrt)] with the packages pscl and emmeans and figures were produced using the package ggplot2 (44). SAS statistical software Version 9.4 was used to conduct the other analyses.

## Results

### Study infants

Of 122 infants screened, 120 were enrolled in the study (60 each in the FF and BF groups) and comprised the FAS population. The details of the FAS and PPS populations are provided in ***Figure 1*** which presents the infant disposition flowchart. A total of 111 infants including 51 FF and 60 BF infants completed the study.

**Figure 1.**
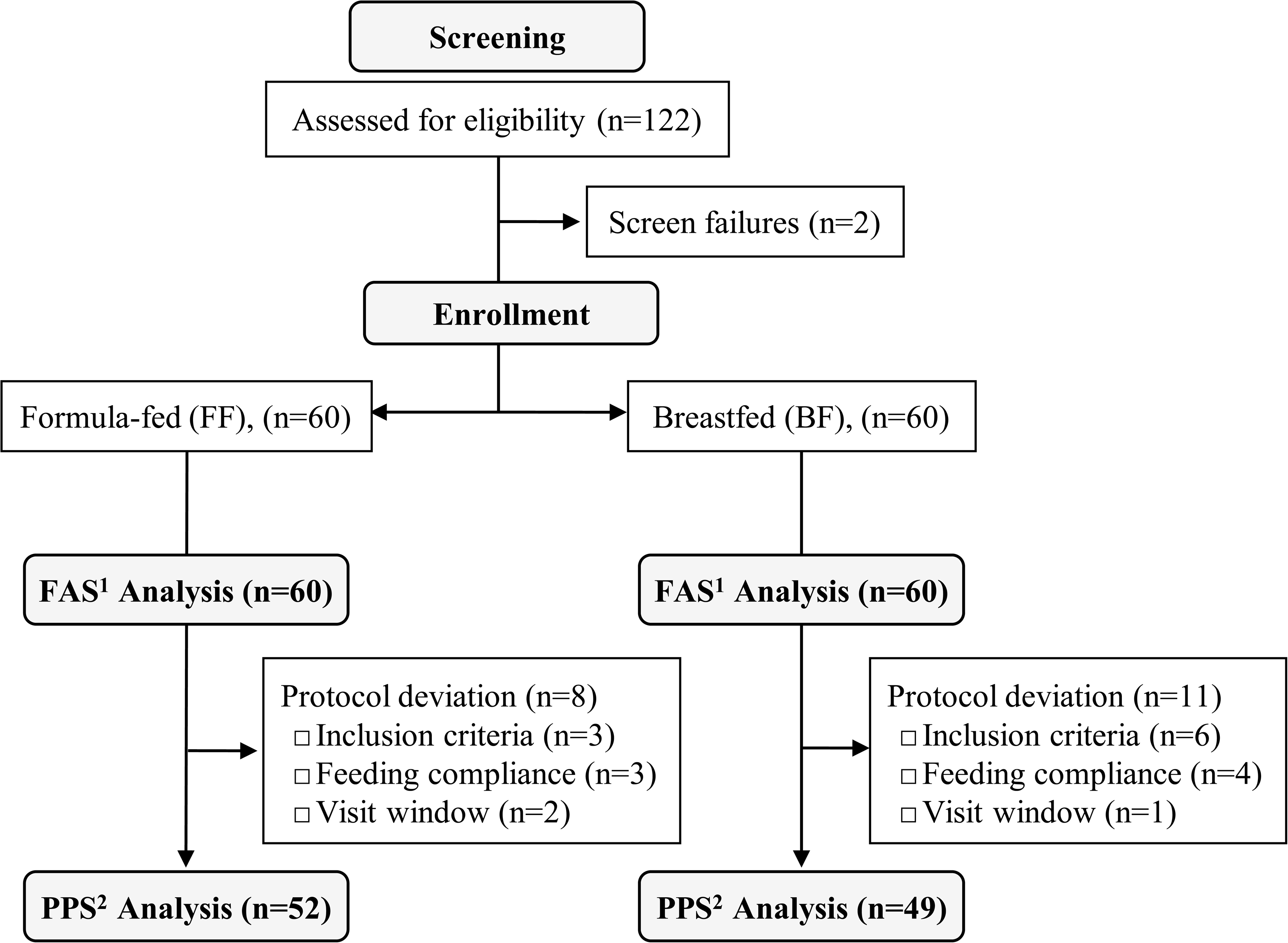
Infant disposition flowchart. ^1^The full analysis set (FAS) includes all infants enrolled who received at least one study feeding (study formula in FF group; breastmilk in BF group). ^2^The per protocol set (PPS) was a subset of the FAS and comprised of infants without any major protocol deviations.

### Baseline demographics and household characteristics

Infant demographics, anthropometrics and household characteristics at baseline were similar between groups except for delivery method (***Table 2).*** A significantly greater proportion of infants in the FF group compared to the BF group were born by cesarean section (55.0% FF, 30.0% BF, p=0.006). In the FF group, 29 (48.3%) infants were exclusively FF prior to enrollment while 31 (51.7%) were predominantly FF. In the BF group, there were 41 (68.3%) and 19 (31.7%) infants who were exclusively and predominantly BF prior to enrollment, respectively. The majority of mothers had completed junior college or a higher level of education (86.7% FF, 83.4% BF).

**Table 2.** Baseline demographic and household characteristics for the full analysis set^1^.

| Characteristic | Total,<br>n=120 | FF, n=60 | BF, n=60 | P-value<sup>2</sup> |
| --- | --- | --- | --- | --- |
| ***Infant demographics*** |  |  |  |  |
| Age at baseline, days | 16.8 (7.8) | 15.5 (8.9) | 18.1 (6.4) | 0.121<sup>b</sup> |
| Gestational age, days | 275.0 (6.9) | 274.8 (6.7) | 275.2 (7.1) | 0.752<sup>a</sup> |
| Birth weight, kg | 3.4 (0.40) | 3.4 (0.36) | 3.4 (0.42) | 0.804<sup>a</sup> |
| Birth length, cm | 49.7 (1.5) | 49.7 (1.4) | 49.8 (1.5) | 0.330<sup>b</sup> |
| Birth head circumference, cm | 34.8 (0.8) | 34.9 (0.7) | 34.6 (0.9) | 0.377<sup>a</sup> |
| Infant sex, % female | 48.3% | 46.7% | 50.0% | 0.715<sup>c</sup> |
| Singleton birth, % yes | 100% | 100% | 100% | - |
| **Type of delivery, % C-section** | **42.5%** | **55.0%** | **30.0%** | **0.006<sup>c</sup>** |
| APGAR at 5 minutes | 10.0 (0.2) | 10.0 (0.13) | 10.0 (0.22) | 0.315<sup>b</sup> |
| ***Household characteristics*** |  |  |  |  |
| Daycare attendance, % No | 100% | 100% | 100% | - |
| Number of children in household | 1.3 (0.5) | 1.3 (0.5) | 1.3 (0.6) | 0.955<sup>b</sup> |
| Maternal education, % completed junior college or higher | 85.0% | 86.7% | 83.4% | 0.480<sup>d</sup> |
| Mother smokes, % no | 100% | 100% | 100% | - |
| Smoking in household, % no | 90.8% | 93.3% | 88.3% | 0.343<sup>c</sup> |

<sup>1</sup>Data presented are mean (SD) unless otherwise specified. The superscript a, b, c, and d of P-values indicate t-test, Wilcoxon rank sum test, Chi-square test and Fisher exact test, respectively.
<sup>2</sup>p-value resulting from two-sided test of difference between the FF and BF groups.

### GI tolerance including GI symptoms and GI-related behaviors

IGSQ index scores demonstrated good GI tolerance in both groups at V2 in the PPS, with mean±SD of 19.9±7.4 for the FF infants and 16.8±4.2 for the BF infants and were not significantly different ***(Figure 2).*** At V2 in the FF Group, 36 infants (76.6%) were considered to have “No GI distress”, 6 infants (12.8%) had “Certain GI distress”, and 5 infants (10.6%) had “Clinically meaningful GI distress”. In the BF Group at V2, 44 infants (89.8%) were considered to have “No GI distress”, 4 infants (8.2%) had “Certain GI distress”, and one infant (2.0%) had “Clinically meaningful GI distress”.

**Figure 2.**
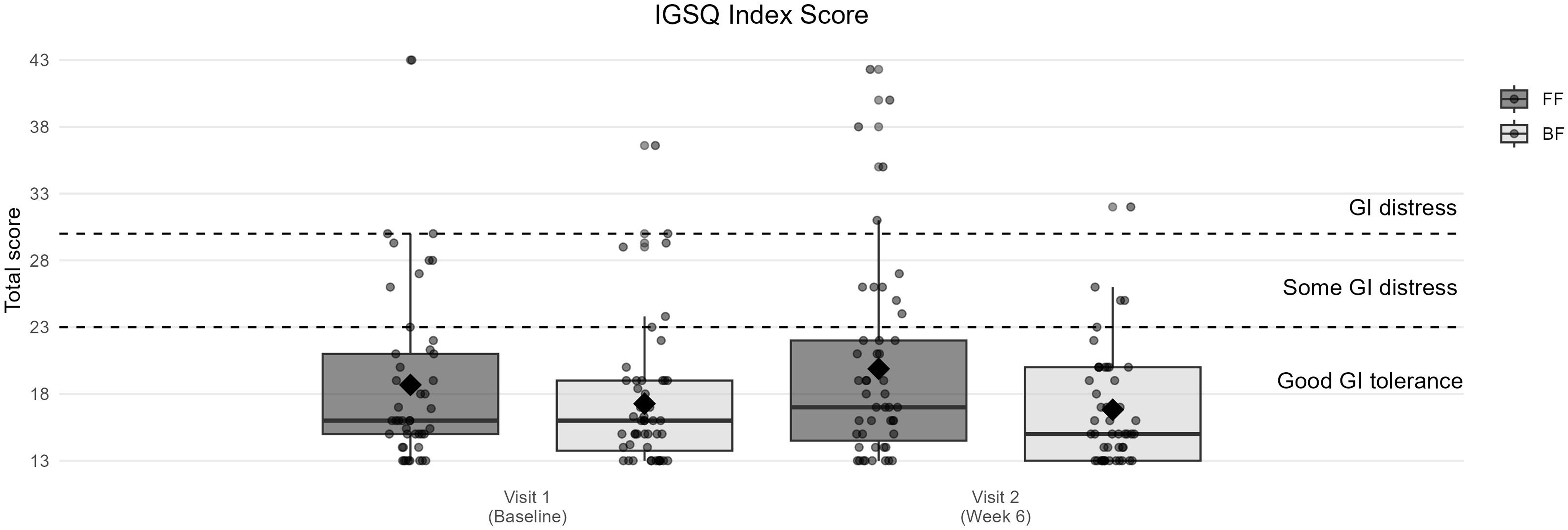
Mean Infant Gastrointestinal Symptom Questionnaire (IGSQ) scores for the per protocol set. The IGSQ index score is calculated from the IGSQ questionnaire and ranges from 13 to 65. From 13 to 23 is good tolerability, > 23 to 30 is problematic or some GI distress, and > 30 to 65 is not well tolerated or GI distress. Data presented are mean (SD) and analyzed by ANCOVA, correcting for the corresponding site, baseline score, delivery mode, sex, and baseline age.

When analyzed by ANCOVA, non-inferiority was not demonstrated in the PPS as the upper bound of the 95% CI was greater than 3.0 (mean difference 1.35 [95% CI: -1.312, 4.012]). However, when analyzed by robust ANCOVA (down weighting the outliers), IGSQ index scores in FF infants were non-inferior to BF infants at V2, with a mean difference 0.90 (95% CI: -0.998, 2.803). Similar results were observed in the FAS.

The IGSQ domain scores for GI symptoms including spitting up/vomiting, flatulence, crying, fussiness and stooling were not significantly different between the FF and BF groups in the FAS at V2 (***Table 3)*.**

**Table 3.** Infant Gastrointestinal Symptom Questionnaire (IGSQ) individual domain scores for the full analysis set^1^.

| ISGQ domain | FF, n=60 |  | BF, n=60 |  | P-value<sup>2</sup> |
| --- | --- | --- | --- | --- | --- |
|  | V1 | V2 | V1 | V2 |  |
| Spitting up / Vomiting | 5.5 (2.3) | 5.6 (2.2) | 5.3 (1.8) | 5.2 (1.7) | 0.185 |
| Flatulence | 3.9 (1.7) | 4.4 (2.0) | 3.9 (1.9) | 3.5 (1.8) | 0.122 |
| Crying | 4.0 (1.7) | 4.2 (2.2) | 3.8 (1.7) | 3.7 (1.1) | 0.611 |
| Fussiness | 2.5 (1.3) | 2.8 (1.8) | 2.4 (1.2) | 2.3 (0.9) | 0.627 |
| Stooling | 2.8 (1.7) | 2.4 (1.0) | 2.1 (0.3) | 2.2 (0.8) | 0.538 |

<sup>1</sup>Data presented are mean (SD) unless otherwise specified.
<sup>2</sup>p-value resulting from a two-sided test of difference between the FF and BF groups estimated with robust ANCOVA.

### Stooling patterns

Stool consistency at V2 was similar across the FF and BF groups, with mean±SD scores of 2.2±0.5 and 2.1±0.5, respectively. Overall, stool consistency in the FF group moved in the direction from mushy/soft to runny (V1 and V2: 39.7% vs 26.4% mushy/soft; 50.0% vs 67.6% runny) which was the predominant stool consistency category in the BF group (V1 and V2: 61.0% vs 72.7% runny). Difficulty in passing stool was low at V1 and V2 in both FF (V1: 1.7%, V2: 6.3%) and BF groups (V1: 1.7%, V2: 1.5%). The incidence of difficulty passing stool was not significantly different at V2 between the FF and BF groups with mean of 7.56% and 6.11%, respectively. Stool frequency was significantly lower in the FF group compared to the BF group at V2 (p<0.001), with the mean±SD number of stools per day being 4.2±2.9 and 7.0±5.4 for the FF and BF groups, respectively.

### Infant quality of life

There were no significant differences in the total score of the IQI between the FF and BF groups at V2 and in change from V1 to V2 (***Additional file 5***). Over 90% of the FF infants slept well and 100% were normally fed and breathing. The majority of FF infants reported normal stool (>88%), were happy/content (>96%), had normal skin (>95%), and were playful/interactive (>98%). No significant differences were observed between the FF and BF groups at V2 nor in change from V1 to V2 for any of the domains including sleeping, feeding, breathing, stooling, mood, skin, and interaction.

### Growth

None of the standardized anthropometric variables, including weight-for-age, length-for-age, weight-for-length, head circumference-for-age, and BMI-for-age z-scores, were significantly different between the FF and BF groups at V2 or in change from V1 to V2 (***Additional file 6***).

### Adverse events and medication use

Physician-reported AEs were comparable in terms of incidence, seriousness, and severity between FF and BF infants ***(Table 4).*** Among 120 total infants, 7 infants had at least one AE in the FF group and 5 infants in the BF group. Serious AEs were slightly higher in the BF compared to the FF group but this difference was not significant (FF: 0%, BF: 6.7%). Both groups had similar mild (FF: 10%, BF: 8.3%) and moderate (FF: 1.7%, BF: 1.7%) AEs. The incidence of common illnesses of interest including acute gastroenteritis/infectious diarrhea (FF: 3.3%, BF: 0.0%), lower respiratory tract infections (FF: 0.0%, BF: 3.3%), and upper respiratory tract infections (FF: 1.7%, BF: 1.7%) were low and not significantly different between the FF and BF groups. Concomitant medications were reported by 2 (3.3%) and 5 (8.3%) infants in the FF and BF groups, respectively. Among these, antibiotic use was reported by one infant in the FF group and 2 in the BF infants and incidence was not significantly different. These infants used cephalosporin antibiotics.

**Table 4.** Physician reported adverse events (AEs), common illnesses, and medication usage for safety analysis set.

|  | FF | BF | P-value |
| --- | --- | --- | --- |
| ***AE reporting*** |  |  |  |
| All AEs, N (%) | 7 (11.7%) | 5 (8.3%) | 0.543<sup>1</sup> |
| Serious AEs, N (%) | 0 (0.0%) | 4 (6.7%) | 0.119<sup>2</sup> |
| Severe AEs, N (%) | 0 (0.0%) | 0 (0.0%) | - |
| Deaths | 0 (0.0%) | 0 (0.0%) | - |
| ***Common illnesses of interest*** |  |  |  |
| Any common illness of interest | 3 (5.0%) | 3 (5.0%) | 1.000<sup>2</sup> |
| Acute gastroenteritis/ infectious diarrhea | 2 (3.3%) | 0 (0.0%) | 0.496<sup>2</sup> |
| Lower respiratory tract infections | 0 (0.0%) | 2 (3.3%) | 0.496<sup>2</sup> |
| Upper respiratory tract infections | 1 (1.7%) | 1 (1.7%) | 1.000<sup>2</sup> |
| ***Medication use*** |  |  |  |
| At least one concomitant medication | 2 (3.3%) | 5 (8.3%) | 0.439<sup>2</sup> |
| Antibiotic use | 1 (1.7%) | 2 (3.3%) | 1.000<sup>2</sup> |

<sup>1</sup>p-value resulting from a two-sided Chi-square test of difference between the FF and BF groups.
<sup>2</sup>p-value resulting from a two-sided Fisher exact test of difference between the FF and BF groups.
N (%) in this table represents the number and percentage of subjects who reported at least one adverse event/common illness of interest/medication use.

### Fecal microbiota composition

At baseline, Shannon diversity was not significantly different between the feeding groups (***Figure 3***). From V1 to V2, there was no significant change in the Shannon diversity in the BF group but a significant change for the FF group (p<0.001). Shannon diversity was significantly different between the feeding groups at V2 (p<0.005).

**Figure 3.**
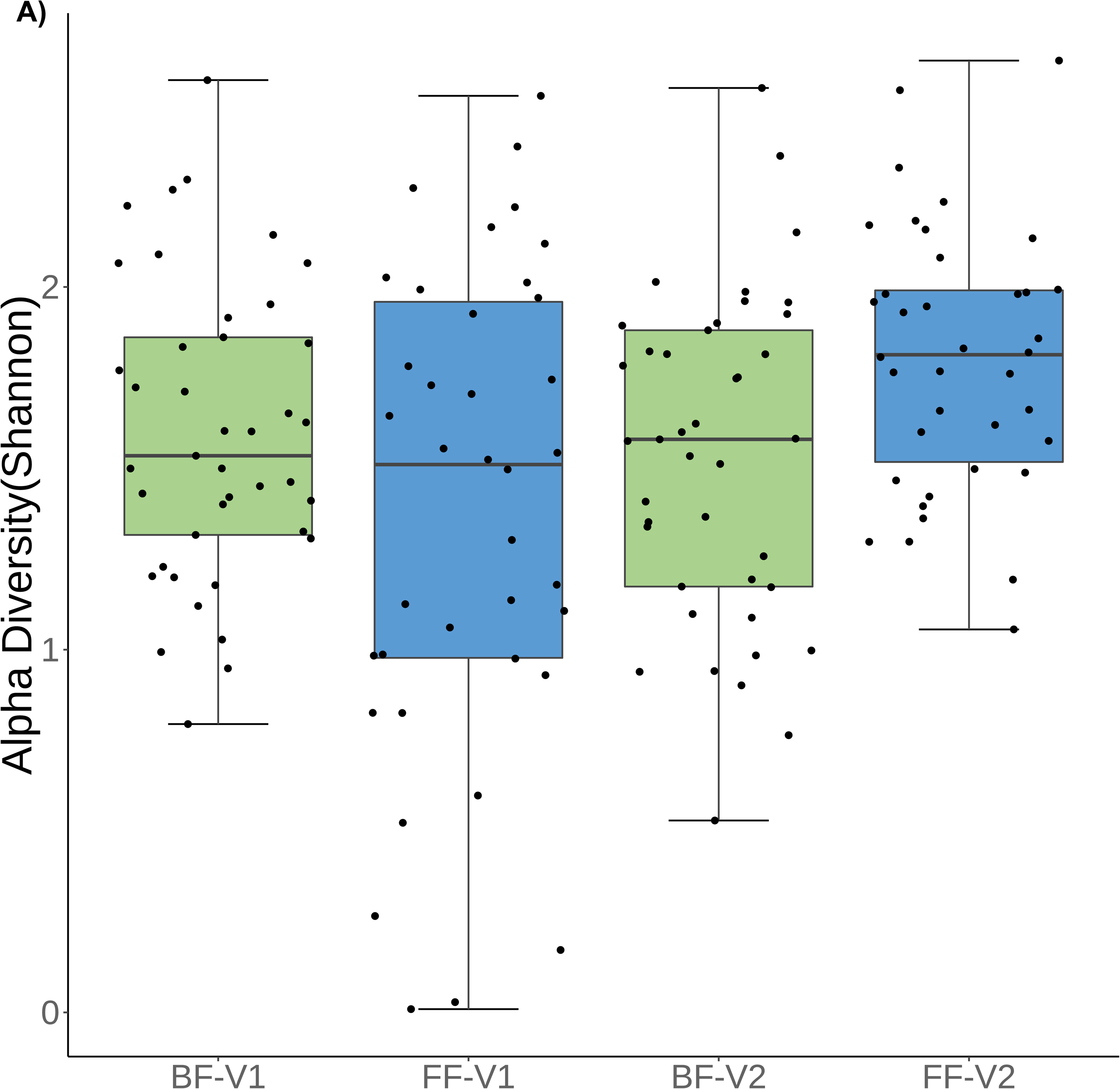
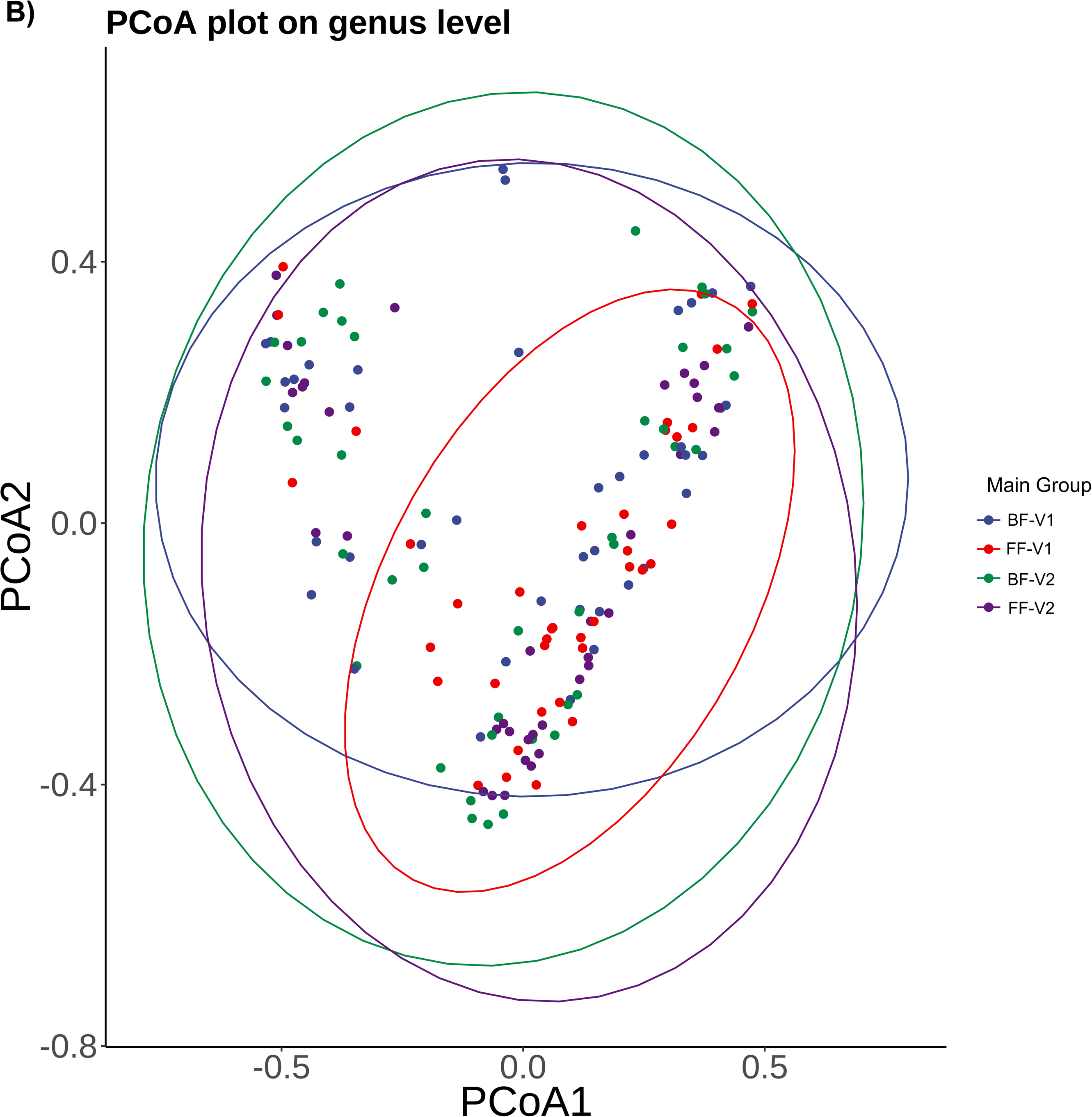
Microbiota modulation with formula intervention after 6 weeks. A) Alpha diversity (Shannon index); B) Beta diversity (PCoA plot).

The fecal community dissimilarities among the 4 study groups defined by feeding regimen and visit (BF-V1, FF-V1, BF-V2, FF-V2) were displayed using PCoA on Bray-Curtis distances calculated at Family, Genus, and Species level. At Genus level (***Figure 3***), FF were significantly different from BF at V1 (p=0.024). From V1 to V2, there were no significant changes in the BF group, and a borderline significant change in the FF group (p=0.054). The FF group was significantly different from the BF group at V2 (p=0.041). Similar trends were observed at Family level (***Additional file 7***) (FF vs. BF at V1: p=0.038; BF at V2 vs. V1: p=0.313; FF at V2 vs. V1: p= 0.023; FF vs. BF at V2: p=0.029) and at Species level (***Additional file 7***) (FF vs. BF at V1: p=0.051; BF at V2 vs. V1: p=0.358; FF at V2 vs. V1: p=0.026; FF vs BF at V2: p=0.034).

### Bifidobacteria

Genus *Bifidobacterium* and seven species of *Bifidobacteria*, which included *Bifidobacterium dentium*, *Bifidobacterium breve*, *Bifidobacterium animalis*, *Bifidobacterium bifidum*, *Bifidobacterium pseudocatenulatum*, *Bifidobacterium adolescentis*, and *Bifidobacterium longum*, were detected in the samples of this study.

There were no significant differences in the relative abundance of the *Bifidobacterium* genus between the FF and BF groups at V2 (p=0.570). Further, the changes in relative abundance of *Bifidobacterium* genus (from V1 to V2) in the FF group were not significantly different from the BF group (p=0.281).

### Fecal metabolites

The total SCFAs and total BCFAs concentration were significantly higher in the FF versus BF group at V2 (p=0.049 and p=0.001 respectively) (***Table 5***). Propionate (p=0.002), valerate (p=0.042), isobutyrate (p=0.002), and isovalerate (p=0.001) were significantly higher in the FF group at V2, but no significant differences were demonstrated for the remaining acids (***Table 5***). Furthermore, the relative proportion of acetate at V2 was significantly lower in the FF group compared to the BF group (p=0.026) (***Additional file 8***). In contrast, the relative proportion of propionate (p=0.012) and isovalerate (p=0.001) at V2 were significantly higher in the FF group. The relative proportion of total BCFAs was also significantly higher in the FF group compared to the BF group (p=0.048). No significant differences were seen between groups in the relative proportions of the remaining acids. Finally, the concentration of fecal metabolites (total SCFAs, total BCFAs, or individual SCFAs) were neither significantly different between vaginal and C-section born infants nor between exclusive and predominant feeding modes (data not shown).

**Table 5.** Levels of SCFAs and BCFAs (μmol/g dry feces) for the full analysis set^1^.

| Metabolite<br>($\mu\text{mol/g dry feces}$) | FF (n=60) | | BF (n=60) | | P-value |
| --- | --- | --- | --- | --- | --- |
|  | V1 | V2 | V1 | V2 | V2 |
| **Total SCFAs** | **109.0 (1.9)** | **164.5 (1.8)** | **58.1 (2.2)** | **87.8 (2.5)** | **0.049** |
| **Total BCFAs** | **0.2 (6.1)** | **0.7 (7.5)** | **0.1 (4.2)** | **0.2 (4.5)** | **0.001** |
| Acetate | 87.3 (1.9) | 122.3 (1.9) | 48.2 (2.1) | 73.2 (2.6) | 0.168 |
| **Propionate** | **3.7 (8.8)** | **16.5 (3.6)** | **2.5 (4.9)** | **4.1 (4.8)** | **0.002** |
| Butyrate | 3.0 (8.4) | 7.4 (5.1) | 2.2 (6.4) | 2.4 (5.3) | 0.147 |
| **Valerate** | **0.07 (4.9)** | **0.1 (4.1)** | **0.04 (3.3)** | **0.05 (3.4)** | **0.042** |
| Caproate | 0.03 (1.8) | 0.03 (2.0) | 0.02 (2.3) | 0.03 (2.2) | 0.801 |
| **Isobutyrate** | **0.1 (5.6)** | **0.3 (7.5)** | **0.06 (4.4)** | **0.1 (4.4)** | **0.002** |
| **Isovalerate** | **0.06 (7.7)** | **0.3 (9.6)** | **0.05 (4.7)** | **0.06 (5.9)** | **0.001** |
| Lactate | 8.4 (3.4) | 12.0 (2.9) | 5.5 (3.6) | 8.4 (3.8) | 0.248 |

BCFA: Branched chain fatty acids; SCFA: Short chain fatty acids
<sup>1</sup>Data presented are geometric mean (geometric SD). SCFAs and BCFAs at V2 were analyzed with ANCOVA models correcting for the corresponding site, baseline value, delivery method, sex, and baseline age. Log-transformation was applied to the dependent variable and baseline value for ANCOVA analysis. P-value results from a two-sided test of difference between the FF and BF groups at V2 estimated with ANCOVA.

### Fecal markers of immune response, inflammation, and intestinal barrier integrity

The means of fecal markers for immune response (sIgA), inflammation (calprotectin, IL-1β, IFN-γ, TNF-α) and intestinal barrier integrity (α1 antitrypsin) were not significantly different between FF and BF groups at V2 (***Figure 4 and Additional file 9)***. Only lipocalin-2 levels at V2 were significantly higher in the BF group compared to the FF group (p=0.048). None of the selected markers at V2 were significantly different between vaginal and C-section born infants or between exclusive and predominant feeding modes (data not shown).

**Figure 4.**
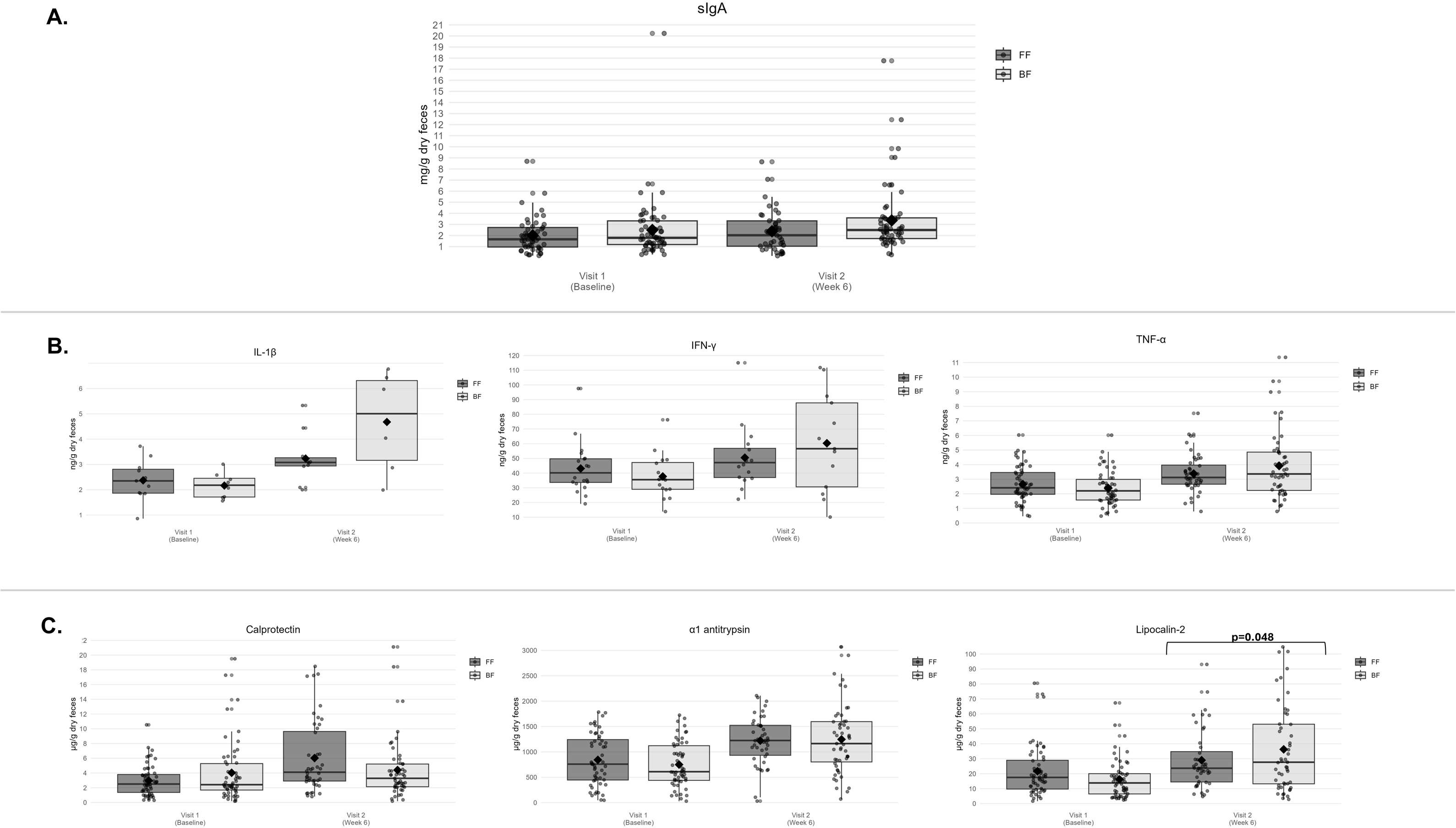
Fecal **A)** Secretory IgA; **B)** Cytokines; and **C)** Intestinal markers for the full analysis set^1^. ^1^Data presented are means (SD). Analyzed by robust ANCOVA models correcting for the corresponding site, baseline value, delivery method, sex, and baseline age.

### Associations of gut microbiota with fecal organic acids

Heatmap of the Spearman correlation coefficients of gut microbiota and SCFAs is reported in ***Figure 5***. At V2, Shannon diversity was significantly associated with propionate (ρ =0.42, FDR p=0.002), with total SCFAs (ρ=0.29, FDR p=0.037) and BCFAs (ρ=0.27, FDR p=0.044).

**Figure 5.**
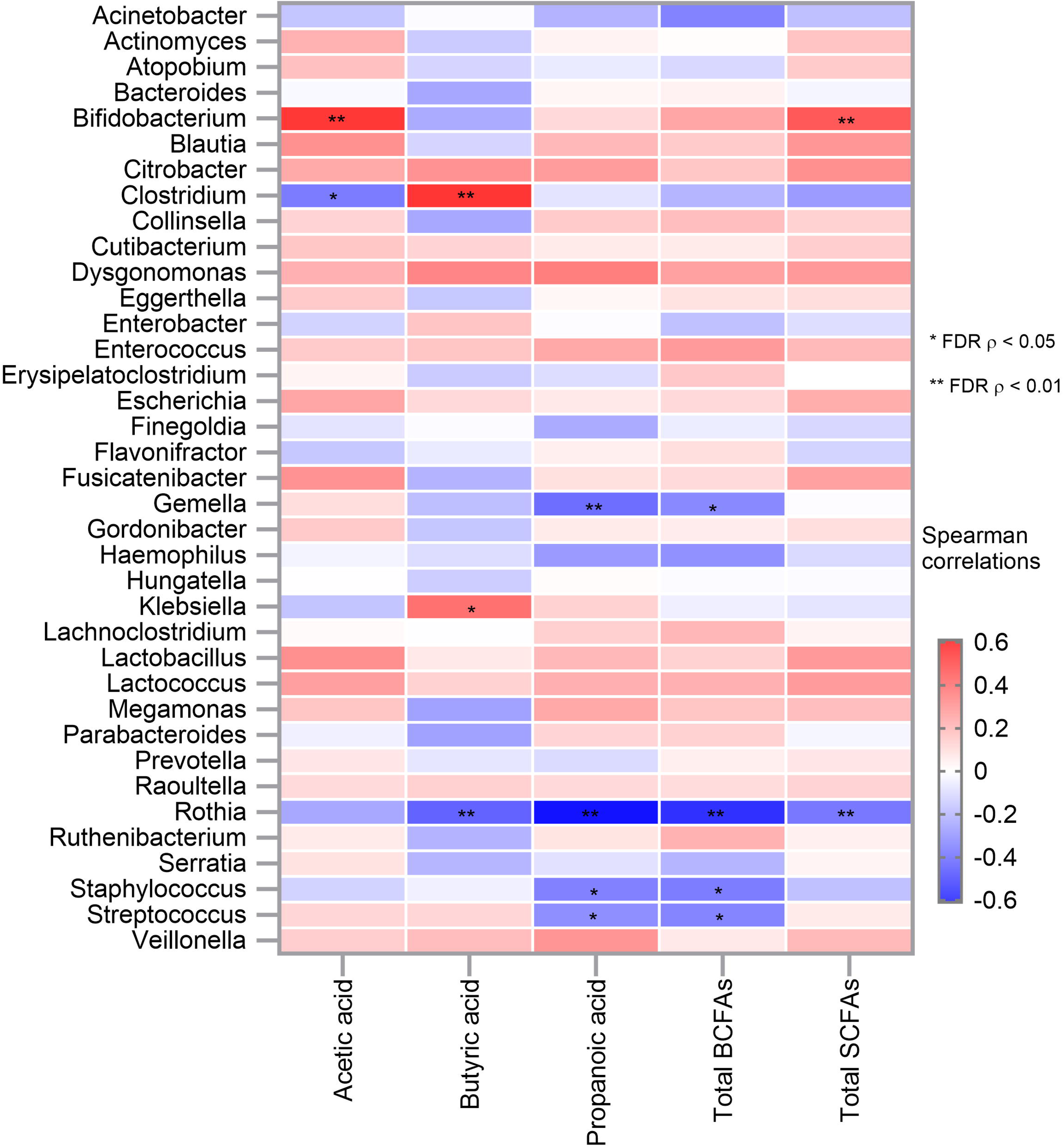
Heatmap of Microbiota vs. SCFAs and BCFAs levels at V2. BCFA: Branched chain fatty acids; SCFA: Short chain fatty acids

Most specifically, at V2 *Bifidobacterium* genus was significantly (FDR p<0.002) and positively associated with total SCFAs and with acetate concentrations, with ρ=0.41 and ρ=0.50 for total SCFAs and acetate respectively. At the species level, these associations were mainly driven by *Bifidobacterium longum* (ρ=0.46, FDR p< 0.0005 and ρ=0.52, FDR p<0.0005 for total SCFAs and acetate, respectively) and by *Bifidobacterium bifidum* (ρ=0.37, FDR p=0.012 for acetate and ρ=0.31, FDR p=0.035 for total SCFAs). Of note, *Bifidobacterium longum* was also significantly associated with total BCFAs (ρ=0.35, FDR p= 0.008).

Interestingly, at V2, mainly significant and negative associations were observed between fecal propionate concentration and gut microbiota such as with *Rothia (*ρ=-0.60, FDR p<0.0001)*, Gemella (*ρ=-0.36, FDR p=0.007), *Staphylococcus (*ρ=-0.32, FDR p=0.026) *and Streptococcus (*ρ=-0.27, FDR p=0.034). Overall, *Rothia* was negatively and significantly associated with most of the main SCFAs (e.g., propionate, butyrate, total BCFAs, total SCFAs) except with acetate (additional Table 10).

## Discussion

The study examined the tolerance and effectiveness of an IF as typically fed outside of a highly controlled clinical trial setting. The age of the infants captures a window period of dynamic change in gut microbiota, establishing a healthy intestinal and immune maturation which can be influenced by nutrition (45, 46). For infants that cannot be breastfed, IF is a suitable alternative that ideally provides adequate nutrition and bioactive components that support the benefits provided by HM. Our results suggest that this IF, with protein and lipid fractions found in HM, is well-tolerated and supports outcomes similar to BF infants.

GI tolerance was examined using the IGSQ which demonstrated good GI tolerance among FF infants similar to that of BF infants. Although non-inferiority was not demonstrated in the primary analyses, GI tolerance in FF infants was non-inferior in sensitivity analyses. Signs of feeding intolerance including GI symptoms such as infrequent or hard stooling, constipation, regurgitation, and colic are often seen in FF infants in the first months of life (47). Depending on the formula composition, stool characteristics often differ between FF and BF infants. FF infants typically have harder stools than BF infants (36, 48), with BF infants typically having softer, loose and more liquid stools (36, 49). In the current study, the overall stool consistency of the FF group moved from mushy/soft to runny which was the predominant stool consistency category of the BF reference group. Components in the study formula has been shown to improve GI tolerance based on previous evidence. IF enriched with α-lactalbumin was shown to improve GI tolerance compared to a control formula and was closer to a BF reference group (23). Similarly, a growing up milk with A2 β-casein when fed to Chinese toddlers with minor GI distress resulted in improved digestive comfort and GI-related symptoms (50).

In addition to good GI tolerance, infants consuming the study formula demonstrated age-appropriate growth similar growth to the BF reference group and tracking close to the median of the WHO growth charts for weight-for-age, length-for-age, weight-for-length, head-circumference-for-age, and BMI-for-age z-scores. These findings are consistent with other studies that have examined IF enriched with specific bioactive compounds such as MFGM (51), α-lactalbumin (52, 53), α-lactalbumin and structured lipids (lipid blend enriched in *Sn-2* palmitate) (54), structured lipids and OF (47), and gangliosides (55). In relation to the rate of adverse events in the current study, it was low and with no significant differences between the FF and BF groups. No safety concerns were identified as reflected by the absence of relevant differences in number or severity of adverse events reported.

Promoting the maturation and function of the intestinal immune system is important for infants to reduce vulnerability to infections and illness (56, 57). Two essential components of the host’s immune development are the GI barrier and GI microbiome (58–60). During early infancy, the diversity and composition of the gut microbial community develop rapidly and are essential for infant development (61). *Bifidobacterium* is one of the initial colonizers of the infant GI tract with important functions in infant development (62, 63). A RCT with an IF containing a lipid blend enriched in *Sn-2 palmitate* showed an increase in *Bifidobacterium* following 8 weeks and an increase in alpha-diversity and acetate at both 8 and 16 weeks (64). IF enriched in MFGM has also been reported to exert bifidogenic effects and increase alpha diversity (65, 66). In this study, the relative abundance of the *Bifidobacterium* genus in the FF group evolved similar to that in the BF reference group.

The infant gut benefits from colonization of bacteria (such as *Bifidobacteria*) that contribute to the production of SCFAs that support metabolism, immunity, and metabolic health (67, 68). In this study, higher levels of SCFAs in stool observed in FF infants may be a consequence of greater bacterial diversity in these infants as reported by the positive association observed between total SCFAs and the Shannon diversity. Of interest, with this IF similar absolute levels of acetate and butyrate in BF and FF infants were observed while others have reported that exclusively BF infants (vs FF infants) showed lower absolute concentrations of acetate, butyrate and propionate (69, 70). This observation suggests that in the study the level of these functional metabolites produced by the gut microbiota were only mildly affected in the FF infants.

Normal composition and vitality of the gut microbiome are specific signs of GI health (58). FF infants often show a different pattern of gut microbial colonization and immune development versus BF infants (71). The current IF supported similar levels in markers of immunity, inflammation, and intestinal barrier function in FF infants compared to BF infants. In the intestinal tract, sIgA plays a crucial role in protecting the intestinal mucosa and regulating host-commensal homeostasis (72, 73). The intestinal production of sIgA starts at approximately one month of age, hence typically, FF infants have lower levels of fecal sIgA in the first months of life compared to BF infants due to the lack of passive transfer from breastmilk (73–76). In the current study, fecal sIgA levels increased from baseline to the end of intervention with no significant difference observed between FF and the BF groups after 6 weeks of intervention (∼ 1.5 to 2.5 months-old infants). These findings are in line with findings from a RCT of IF supplemented with osteopontin which resulted in immune outcomes such as cytokine response closer to that of BF infants (27).

Breastfeeding is also associated with lower risk of respiratory and GI infections (77). Interestingly, in the current study, there were no significant differences in common illnesses of interest between the FF and BF groups.

Several factors positively impact GI tolerance and intestinal health which in turn support infant quality of life. In this study, infant quality of life including sleeping, feeding, breathing, stooling, mood, skin, and interaction was comparable between the FF and BF groups. The quality of life scores in our study are in line with high quality of life scores observed in previous studies on IF enriched with α-lactalbumin (78) and α-lactalbumin, *Sn-2,* and OF (79).

Strengths of our study include the high infant completion rate, particularly during the COVID-19 era, which minimizes potential for bias due to loss to follow-up and missing data. Also, the use of a validated tool to assess GI symptoms and pictorial presentation for stools to reduce the chances of mischaracterization of stool consistency parameters. We strengthened the assessment of the impact of the IF on infant intestinal and immune health via examination of the microbiota and metabolites produced, as well as selected fecal biomarkers. While it could have been interesting to analyze systemic immunity markers, infant blood collection is challenging and was not included in this study. Another consideration in our study was the recruitment of predominantly FF infants (>75% of total milk feeds per day) and not exclusively FF infants. While 25% of the intake may be coming from other sources including HM, this reflects the reality of infant feeding practices (80).

There are some limitations in our study. One limitation is the short study duration not allowing to assess long-term effects. Further, a single arm study design may bias the results, but this was minimized by inclusion of a gold standard BF reference group as comparator. Due to the self-selection or predetermined nature of feeding groups, there may be potential selection bias or confounding variables (e.g., socio-economic status, maternal health, and baseline microbiome differences) that may affect the results. While the IGSQ is a scientifically designed, standardized and validated tool for obtaining parents’ perspectives on the frequency and severity of infants’ GI signs and symptoms and provides information that is interpretable and meaningful to practicing clinicians, it is subjective in nature.

In conclusion, while HM is considered the optimal source of nutrition for infants, advancements in IF composition are needed to offer the best possible alternative for infants who cannot be fed HM. Consumption of an IF which has a whey protein profile that delivers a formula enriched in α-lactalbumin, osteopontin, lactoferrin, MFGM proteins and lipids, and a casein profile comprised of A2 β-casein, in combination with an *Sn-2* palmitate enriched TAG core resulted in feeding tolerance, safety, and intestinal and immune health profiles similar to those of BF infants in a Chinese population.

## Supporting information

Addiitonal-files from 1 to 10

## Data Availability

The datasets used and/or analyzed during the current study are available from the corresponding author on reasonable request.

## Acknowledgements

The authors would like to thank the families of all infants who participated in the study and the research staff at each of the study sites. We thank the following people from Tigermed, China: Chang Qi for support on statistical analyses, Zhe Cheng for support on project management at the study sites, Yifei Zhang for data monitoring and Binyi Wu for data management. Thanks to Human Metabolomics Institutes, Guangdong, China for analyses of stool biomarkers and Beijing Genomics Institute, Shenzhen, China for analyses of stool microbiota. Our sincere thanks to Manjiang Yao and Colin Cercamondi for their contributions to the study conception, design and set up and Irma Silva-Zolezzi and Manjiang Yao for their critical review and feedback on the manuscript.

## Declarations

### Author Contribution Statement

YW, ML and WC: Funding acquisition, Investigation, Project administration, Resources, Supervision, Writing – review & editing, TMS: Formal analyses, Investigation, Project administration, Resources, Writing – original draft, Writing – review & editing, SKD, KV, JPG: Formal analysis, Investigation, Writing – original draft, Writing – review & editing, JB, EK and JR: Writing – original draft, Writing – review & editing, ND, XW, LR: Data curation, Formal analysis, Visualization, Writing – original draft, Writing – review & editing, QL: Investigation, Project administration, Resources, Writing – review & editing, ATV: Formal analysis, Investigation, Writing –review & editing, JD Writing – review & editing, KW and QZ: Analyses of microbiome samples, Writing – review & editing.

### Ethics approval and consent to participate

This study was approved by the ethics committees at the two institutions in China which were the Ethics Committee of Xinhua Hospital Affiliated to Shanghai Jiao Tong University School of Medicine (XHEC-C-2020-053-1, and the Ethics Committee of Shanghai Public Health Clinical Center (2020-E151-01, respectively and conducted in compliance with the Declaration of Helsinki and the International Conference on Harmonization Guidelines for Good Clinical Practice. This study was also approved by the Human Genetic Resource Administrative Committee of the Ministry of Science and Technology of China according to the applicable regulation before the study was initiated. Prior to enrollment, parent(s)/legally authorized representative provided written informed consent. This trial was registered on ClinicalTrials.gov (NCT04880083).

### Consent for publication

Not applicable as the manuscript does not contain data from any individual person.

## Abbreviations

AE: Adverse event
alpha (α)-lactalbumin: α-lactalbumin
ANCOVA: Analysis of covariance
BCFA: Branched chain fatty acid
BF: Breastfed
BP: Base pair
BMI: Body mass index
CI: Confidence interval
CONSORT: Consolidated Standards of Reporting Trials
DIP-Seq: DNA immunoprecipitation sequencing
DNA: Deoxyribonucleic acid
FAS: Full analysis set
FF: Formula-fed
GI: Gastrointestinal
HM: Human milk
IF: Infant Formula
IFN-γ: Interferon gamma
IGSQ-13: Infant Gastrointestinal Symptom Questionnaire-13
IL-1β: Interleukin-1 beta
IQI: Infant Quality of Life Instrument
LLOQ: Low limit of quantification
LOD: Limit of detection
MetaPhlAn3: Metagenomic Phylogenetic Analysis
MFGM: Milk-fat-globule membrane
OF: Oligofructose
OPO: *Sn-2* palmitate
PCoA: Principal coordinates analysis
PC: Phosphatidylcholine
PE: Phosphatidylethanolamine
PI: Phosphatidylinositol
PL: Phospholipids
PS: Phosphatidylserine
Permanova: Permutational multivariate analysis of variance
PPS: Per protocol set
QoL: Quality of life
RCT: Randomized clinical trial
SAE: Serious adverse events
SCFA: Short chain fatty acid
SD: Standard deviation
sIgA: Secretory immunoglobulin A
SS: Safety set
TAG: Triacylglycerol
TNF-α: Tumor necrosis factor -alpha
ULOQ: Upper limit of quantification
V1: Baseline visit
V2: Study end visit
WHO: World Health Organization
ZINB: Zero-inflated negative binomial

## Additional files

**Additional file 1.** Consolidated Standards of Reporting Trials (CONSORT) Checklist.

**Additional file 2.** Supplementary methods for the microbiota analyses.

**Additional file 3.** LLOQ and ULOQ of fecal SCFAs analysis.

**Additional file 4.** LLOQ and ULOQ of fecal markers of immune response, inflammation, and intestinal barrier integrity.

**Additional file 5.** Infant quality of life for the full analysis set.

**Additional file 6.** Infant anthropometrics for the full analysis set.

**Additional file 7.** A) Principal coordinates analysis (PCoA) plot on species level; B) PCoA plot on family level.

**Additional file 8.** Relative proportion (%) of fecal SCFAs and BCFAs for the full analysis set. **Additional file 9.** Fecal biomarkers of immune response, inflammation, and intestinal barrier integrity for the full analysis set.

**Additional file 10.** Spearman rank correlations between gut microbiota and fecal organic acids at V2 with Benjamini-Hochberg false discovery rate correction applied to the set of two-sided p-values.

