## Supplementary material for "Effects of an infant formula containing a whey protein concentrate on feeding tolerance and markers of intestinal immune defense in Chinese infants": Addiitonal-files from 1 to 10

**Additional file 1.** Consolidated Standards of Reporting Trials (CONSORT) Checklist.

**Additional file 2.** Supplementary methods for the microbiota analyses.

**Additional file 3.** LLOQ and ULOQ of fecal SCFAs analysis.

**Additional file 4.** LLOQ and ULOQ of fecal markers of immune response, inflammation, and intestinal barrier integrity.

**Additional file 5.** Infant quality of life for the full analysis set.

**Additional file 6.** Infant anthropometrics for the full analysis set.

**Additional file 7.** A) Principal coordinates analysis (PCoA) plot on species level; B) PCoA plot on family level.

**Additional file 8.** Relative proportion (%) of fecal SCFAs and BCFAs for the full analysis set.

**Additional file 9.** Fecal biomarkers of immune response, inflammation, and intestinal barrier integrity for the full analysis set.

**Additional file 10.** Spearman rank correlations between gut microbiota and fecal organic acids at visit 2 (V2) with Benjamini-Hochberg false discovery rate correction applied to the set of two-sided p-values.

**Additional file 1.** Consolidated Standards of Reporting Trials (CONSORT) Checklist.

### CONSORT 2010 checklist of information *#

| Section/Topic | Item No | Checklist item | Reported on page No |
|---|---|---|---|
| **Title and abstract** | | | |
| | 1a | Identification as a randomised trial in the title | Not applicable |
| | 1b | Structured summary of trial design, methods, results, and conclusions (for specific guidance see CONSORT for abstracts) | pp.4-5 |
| **Introduction** | | | |
| Background and objectives | 2a | Scientific background and explanation of rationale | pp.6-9 |
| | 2b | Specific objectives or hypotheses | p.10 |
| | | **Methods** | |
| Trial design | 3a | Description of trial design (such as parallel, factorial) including allocation ratio | p.10 |
| | 3b | Important changes to methods after trial commencement (such as eligibility criteria), with reasons | Not applicable |
| Participants | 4a | Eligibility criteria for participants | pp.10-11 |
| | 4b | Settings and locations where the data were collected | p.10 |
| Interventions | 5 | The interventions for each group with sufficient details to allow replication, including how and when they were actually administered | pp.11-12 |
| Outcomes | 6a | Completely defined pre-specified primary and secondary outcome measures, including how and when they were assessed | pp.12-17 |
| | 6b | Any changes to trial outcomes after the trial commenced, with reasons | Not applicable |
| Sample size | 7a | How sample size was determined | pp. 17-18 |
| | 7b | When applicable, explanation of any interim analyses and stopping guidelines | Not applicable |
| Randomisation: | | | Not applicable |
| Sequence generation | 8a | Method used to generate the random allocation sequence | |
| | 8b | Type of randomisation; details of any restriction (such as blocking and block size) | Not applicable; single-arm |
| Allocation concealment mechanism | 9 | Mechanism used to implement the random allocation sequence (such as sequentially numbered containers), describing any steps taken to conceal the sequence until interventions were assigned | Not applicable |

| | | | |
|---|---|---|---|
| Implementation | 10 | Who generated the random allocation sequence, who enrolled participants, and who assigned participants to interventions | Not applicable |
| Blinding | 11a | If done, who was blinded after assignment to interventions (for example, participants, care providers, those assessing outcomes) and how | Not applicable |
| | 11b | If relevant, description of the similarity of interventions | Not applicable |
| Statistical methods | 12a | Statistical methods used to compare groups for primary and secondary outcomes | pp.18-21 |
| | 12b | Methods for additional analyses, such as subgroup analyses and adjusted analyses | pp.18-21 |
| | | **Results** | |
| Participant flow (a diagram is strongly recommended) | 13a | For each group, the numbers of participants who were randomly assigned, received intended treatment, and were analysed for the primary outcome | p.21 |
| | 13b | For each group, losses and exclusions after randomisation, together with reasons | p.21 |
| Recruitment | 14a | Dates defining the periods of recruitment and follow-up | p.10 |
| | 14b | Why the trial ended or was stopped | Not applicable |
| Baseline data | 15 | A table showing baseline demographic and clinical characteristics for each group | p.21-22 |
| Numbers analysed | 16 | For each group, number of participants (denominator) included in each analysis and whether the analysis was by original assigned groups | Tables/Figures/ Additional files/Text pp.21-30 |
| Outcomes and estimation | 17a | For each primary and secondary outcome, results for each group, and the estimated effect size and its precision (such as 95% confidence interval) | Tables/Figures/ Additional files/Text pp.21-30 |
| | 17b | For binary outcomes, presentation of both absolute and relative effect sizes is recommended | Not applicable |
| Ancillary analyses | 18 | Results of any other analyses performed, including subgroup analyses and adjusted analyses, distinguishing pre-specified from exploratory | Tables/Figures/ Additional files/Text pp.21-30 |
| Harms | 19 | All important harms or unintended effects in each group (for specific guidance see CONSORT for harms) | pp.25-26/Table 4 |
| | | **Discussion** | |
| Limitations | 20 | Trial limitations, addressing sources of potential bias, imprecision, and, if relevant, multiplicity of analyses | pp.33-34 |
| Generalisability | 21 | Generalisability (external validity, applicability) of the trial findings | pp.33-34 |

| | | | |
|---|---|---|---|
| Interpretation | 22 | Interpretation consistent with results, balancing benefits and harms, and considering other relevant evidence | pp.30-34 |
| **Other information** | | | |
| Registration | 23 | Registration number and name of trial registry | p.11 |
| Protocol | 24 | Where the full trial protocol can be accessed, if available | Not applicable, trial details at CT.Gov |
| Funding | 25 | Sources of funding and other support (such as supply of drugs), role of funders | p.2 |

Citation: Schulz KF, Altman DG, Moher D, for the CONSORT Group. CONSORT 2010 Statement: updated guidelines for reporting parallel group randomised trials. BMC Medicine. 2010;8:18.

© 2010 Schulz et al. This is an Open Access article distributed under the terms of the Creative Commons Attribution License (http://creativecommons.org/licenses/by/2.0), which permits unrestricted use, distribution, and reproduction in any medium, provided the original work is properly cited.

*We strongly recommend reading this statement in conjunction with the CONSORT 2010 Explanation and Elaboration for important clarifications on all the items. If relevant, we also recommend reading CONSORT extensions for cluster randomised trials, non-inferiority and equivalence trials, non-pharmacological treatments, herbal interventions, and pragmatic trials. Additional extensions are forthcoming: for those and for up-to-date references relevant to this checklist, see www.consort-statement.org.

### Considering that this is a non-randomized study, we have used applicable parts of the guidance to report the trial .

**Additional file 2.** Supplementary methods for the microbiota analyses.

***DNA extraction and sequencing***

Magpure Stool DNA KF Kit B (Magen, China) was used to extract DNA from fecal samples, following the manufacturer's instructions. DNA was fragmented with Covaris E220 (Covaris, Brighton, UK) to yield 300 to 700 bp of fragments. After purification, end-repairing, A-tailing and PCR amplification, PCR products were denatured to generate a single-strand circular DNA library using MGIeasy general DNA library preparation kit (MGI, China) following the manufacturer's instructions. DNA libraries were loaded into a DIPSEQ platform (MGI, China) for sequencing to receive 40 million paired-end 100bp reads per sample.

***Data processing***

BGI sequencing parameter settings (including host genome reference, adaptor sequence etc.) were used. As an analysis tool developed for quality control and preprocessing of FASTQ and SAM/BAM data, SOAPnuke includes 5 modules for different usage scenarios, namely filter, filterHts, filterStLFR, filtersRNA and filterMeta. In this study, we used filterMeta modules for filter raw data. The main commonly used parameters of the software are as follows: –lowQual:low quality threshold, –qualRate:low quality rate, –nRate:N rate threshold,–thread:threads number used in process,–adapter1:adapter sequence or list file of read1,–adapter2:adapter sequence or list file of read2. The parameter we set: SOAPnuke filterMeta –lowQual 15 –qualRate 0.2 –nRate 0.05 –thread 4 –adapter1 AAGTCGGAGGCCAAGCGGTCTTAGGAAGACAA –adapter2 AAGTCGGATCGTAGCCATGTCGTTCTGTGAGCCAAGGAGTTG. The adapter sequence presented above is added when constructing library. During sequencing process, there would be a primer matching the adapter.

SOAP2: an improved ultrafast tool for short read alignment has become a common tool in microbial ecology. However, certain samples especially from host-associated environments can contain a high degree of DNA sequences derived from hosts. In this study, we used SOAP2 alignment of reference genomes hg38 fasta. The parameters of the software are as follows: -r: how to report repeat hits, 0=none; 1=random one; 2=all, -p: number of processors to use, -m: minimal insert size allowed, -x: maximal insert size allowed. We set: -r 1 -p 10 -m 100 -x 1000. MetaPhlAn (version 3) was used to profile the composition of microbial communities (Bacteria, Archaea and Eukaryotes) from metagenomic shotgun sequencing data with species-level.

MetaPhlAn relies on ~1.1M unique clade-specific marker genes identified from ~100,000 reference genomes (~99,500 bacterial and archaeal and ~500 eukaryotic). MetaPhlAn3 was used with default parameters to annotation and profile acquisition of taxons. The script is: metaphlan metagenome.fastq –input_type fastq -o profiled_metagenome.txt. For subsequent analysis, read counts were transformed into relative abundances by normalization to the total number of reads per sample by MetaPhlAn.

***Quality control - Data Quality***

*Sequencing quality*

The sequencing quality was monitored by a variety of indicators such as read coverage, Q30 (The percentage of bases with a quality score of 30 or higher, respectively ), Q20, GC content, % adaptor reads, % human genome contamination. 61 of 166 (97.0%) fecal samples have >=5 GB sequencing raw data size (5 samples with less than 5GB data size). These 5 samples were removed from the following analyses.

Heatmaps were created to visualize the difference between each batch, using both the Pearson correlation and the Spearman correlation. No obvious batch effects were observed.

##### *Possible contamination – High E. coli relative abundance*

In the species-level fecal microbial community profile, we observed that 4 samples had an exorbitant relative abundance of *E. coli*, exceeding 90. It is may due to either environmental contamination or the sampling methodology employed. If we can confidently assert that both the sampling procedures and the environmental conditions were rigorously controlled and maintained, we must consider the possibility that this data accurately reflects the underlying reality. Interesting, the results are consistent with two previous studies (1, 2). These studies reported a similar trend to ours, demonstrating that E. coli's relative abundance is notably elevated during the early stages of infancy, often exceeding 90%, before gradually diminishing as the gut matures. Furthermore, in concurrence with our findings, these studies also observed a higher prevalence of E. coli in infants delivered vaginally. Additionally, those 4 samples all came from FF group at visit 1. Deleting them would impact sample sizes esp. in paired analysis. So, these 4 samples were kept for the subsequent analysis.

##### *Clinical outliers*

Two subjects failed screening and were excluded from the full analysis set (FAS). One subject (s1) had contributed two fecal samples, which were included in the set of 165 samples for sequencing. The other subject (s2) had not contributed any fecal samples. Since the gut microbiota analysis was conducted in the FAS population and s1 was excluded from it, the two microbiome sequencing samples belonging to s1 were not included in the statistical analysis.

##### *Fecal sample description*

Overall, data from 165 samples were obtained using the DNBSEQ platform. 5 sample were removed for failing the minimum sequencing raw data size of 5Gb (as mentioned above). Further 2 samples belonging to a subject (s1), who failed clinical screening, were removed (as mentioned above). The remaining 158 samples were used for the analysis. The table below presents the enrolled subjects by intervention group and reference group of "FAS population after quality control" (158 samples).

Overview of FAS subjects with microbiota data (after QC etc.).

| Visit | Group | Frequency |
|---|---|---|
| V1 | BF | 41 |
| V2 | BF | 41 |
| V1 | FF | 38 |
| V2 | FF | 38 |

**Additional file 3.** LLOQ and ULOQ of fecal SCFAs analysis.

| | **LLOQ (μmol/L)** | **ULOQ (μmol/L)** |
|---|---|---|
| Acetate | 100 | 20000 |
| Propionate | 25 | 5000 |
| Butyrate | 25 | 5000 |
| Valerate | 1 | 200 |
| Isobutyrate | 2.5 | 500 |
| Isovalerate | 1 | 200 |
| Ethylmethylacetate | 1 | 100 |
| Caproate | 0.5 | 100 |
| 4-methylvalerate | 0.5 | 100 |
| 3-hydroxyisovalerate | 0.5 | 100 |
| Lactate | 100 | 5000 |

LLOQ: Lower limit of quantification; SCFA: Short chain fatty acid; ULOQ: Upper limit of quantification.

**Additional file 4.** LLOQ and ULOQ of fecal markers of immune response, inflammation, and intestinal barrier integrity.

| | LLOQ | ULOQ | LOD |
|---|---|---|---|
| sIgA (ng/ml) | 0.31 | 20 | 0.19 |
| Calprotectin (pg/ml) | 37.8 | 3200 | 35 |
| TNF-α (pg/ml) | 2.5 | 80 | 0.1 |
| IL-1β (pg/ml) | 2.5 | 80 | 0.1 |
| IFN-γ (pg/ml) | 25 | 800 | 1.0 |
| Lipocalin-2 (pg/ml) | 46.9 | 3000 | 14.8 |
| α1 antitrypsin (ng/ml) | 0.39 | 25 | 0.31 |

LLOQ: Lower limit of quantification; LOD: Limit of detection; sIgA: Secretory immunoglobulin A; ULOQ: Upper limit of quantification.

**Additional file 5.** Infant quality of life for the full analysis set[1].

| IQI domain | FF (n=60) | | BF (n=60) | | P-value |
|---|---|---|---|---|---|
| | **V1** | **V2** | **V1** | **V2** | |
| Total Score[2] | -0.06 (0.03) | -0.07 (0.06) | -0.07 (0.03) | -0.08 (0.08) | 0.255 |
| Sleeping | | | | | 0.076 |
| Sleeps well | 59 (98.3%) | 49 (94.2%) | 59 (98.3%) | 54 (91.5%) | |
| Slightly affected sleep | 1 (1.7%) | 3 (5.8%) | 1 (1.7%) | 4 (6.8%) | |
| Moderately affected sleep | 0 (0.0%) | 0 (0.0%) | 0 (0.0%) | 1 (1.7%) | |
| Severely disturbed sleep | 0 (0.0%) | 0 (0.0%) | 0 (0.0%) | 0 (0.0%) | |
| Feeding | | | | | 0.152 |
| Normal feeding | 60 (100.0%) | 52 (100.0%) | 60 (100.0 %) | 57 (96.6%) | |
| Slight feeding problems | 0 (0.0%) | 0 (0.0%) | 0 (0.0%) | 2 (3.4%) | |
| Moderate feeding problems | 0 (0.0%) | 0 (0.0%) | 0 (0.0%) | 0 (0.0%) | |
| Severe feeding problems | 0 (0.0%) | 0 (0.0%) | 0 (0.0%) | 0 (0.0%) | |
| Breathing | | | | | 0.221 |
| Normal breathing | 60 (100.0%) | 52 (100.0%) | 60 (100.0%) | 58 (98.3%) | |
| Slight breathing problems | 0 (0.0%) | 0 (0.0%) | 0 (0.0%) | 1 (1.7%) | |
| Moderate breathing problems | 0 (0.0%) | 0 (0.0%) | 0 (0.0%) | 0 (0.0%) | |
| Severe breathing problems | 0 (0.0%) | 0 (0.0%) | 0 (0.0%) | 0 (0.0%) | |
| Stooling | | | | | 0.253 |
| Normal stool | 60 (100.0%) | 46 (88.5%) | 58 (96.7%) | 58 (98.3%) | |
| Slight stool problems | 0 (0.0%) | 5 (9.6%) | 2 (3.3%) | 1 (1.7%) | |
| Moderate stool problems | 0 (0.0%) | 1 (1.9%) | 0 (0.0%) | 0 (0.0%) | |
| Severe stool problems | 0 (0.0%) | 0 (0.0%) | 0 (0.0%) | 0 (0.0%) | |

| | | | | | |
|---|---|---|---|---|---|
| Mood | | | | | 0.977 |
| Happy/content | 60 (100.0%) | 50 (96.2%) | 60 (100.0%) | 57 (96.6%) | |
| Fussy/irritable | 0 (0.0%) | 0 (0.0%) | 0 (0.0%) | 0 (0.0%) | |
| Crying | 0 (0.0%) | 2 (3.8%) | 0 (0.0%) | 2 (3.4%) | |
| Inconsolable crying | 0 (0.0%) | 0 (0.0%) | 0 (0.0%) | 0 (0.0%) | |
| Skin | | | | | 0.089 |
| Normal skin | 57 (95.0%) | 50 (96.2%) | 57 (95.0%) | 54 (91.5%) | |
| Dry or red skin | 2 (3.3%) | 2 (3.8%) | 3 (5.0%) | 5 (8.5%) | |
| Irritated or itchy skin | 1 (1.7%) | 0 (0.0%) | 0 (0.0%) | 0 (0.0%) | |
| Bleeding or cracked skin | 0 (0.0%) | 0 (0.0%) | 0 (0.0%) | 0 (0.0%) | |
| Interaction | | | | | 0.912 |
| Highly playful/interactive | 51 (85.0%) | 43 (82.7%) | 53 (88.3%) | 50 (84.7%) | |
| Playful/interactive | 9 (15.0%) | 8 (15.4%) | 7 (11.7%) | 9 (15.3%) | |
| Less playful/less interactive | 0 (0.0%) | 1 (1.9%) | 0 (0.0%) | 0 (0.0%) | |
| Low-energy/inactive/dull | 0 (0.0%) | 0 (0.0%) | 0 (0.0%) | 0 (0.0%) | |

BF: Breastfed; FF: Formula-fed; IQI: Infant Quality of Life Questionnaire; V1: Visit 1; V2: Visit 2.

[1]Data presented are N (%). The total IQI score ranges from 0 to -1.12, with 0 indicating no symptoms and -1.12 indicating the worst symptoms. Categorical responses were scored from 1 to 4 for each item (total of 7 items related to breathing, mood, sleeping, feeding, skin, interaction, and stooling), with increasing scores indicating problematic states. Continuous scores were analyzed by ANCOVA models correcting for the corresponding site, baseline value, delivery method, sex, and baseline age. P-value results from a two-sided test of difference between the FF and BF groups at V2 estimated with ANCOVA.

[2] Data presented are mean (SD).

**Additional file 6.** Infant anthropometrics for the full analysis set.

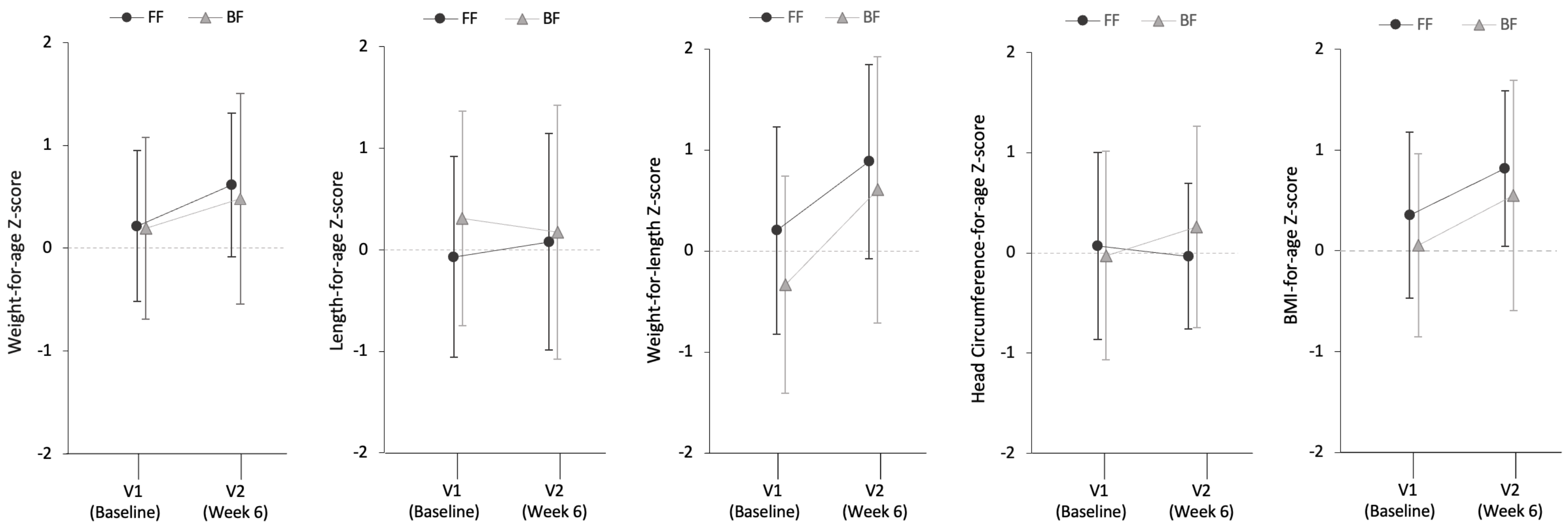


BF: Breastfed; FF: Formula-fed; V1: Visit 1; V2: Visit 2. There were no significant differences between groups at V2 (week 6) for weight-for-age (p=0.27), length-for-age (p=0.38), weight-for-length (p=0.71), head circumference-for-age (p=0.36), and BMI-for-age z-scores (p=0.38).

**Additional file 7.** A) Principal coordinates analysis (PCoA) plot on species level; B) PCoA plot on family level*.

**A)**

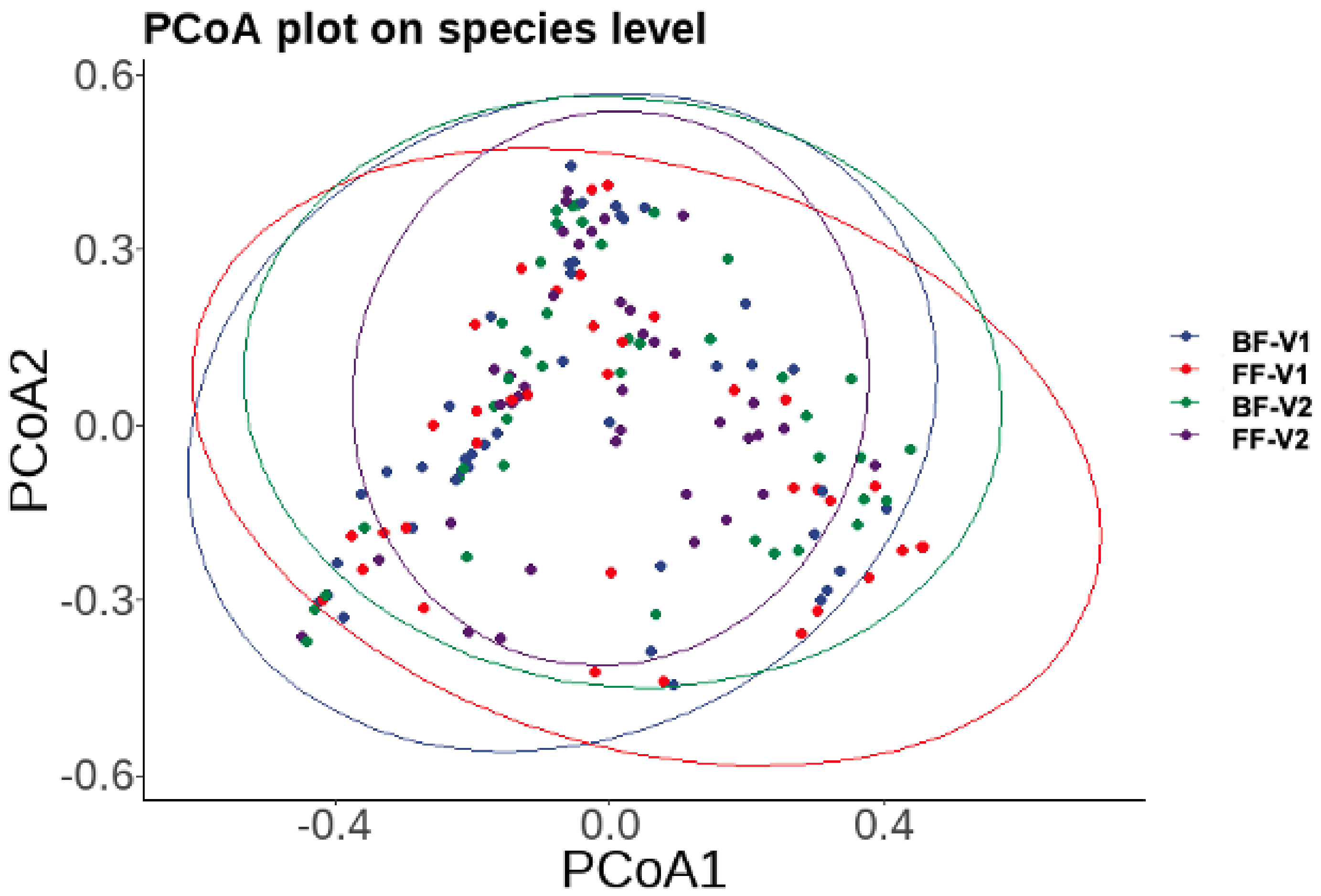


*Points of different colors represent samples of different groups.

B)

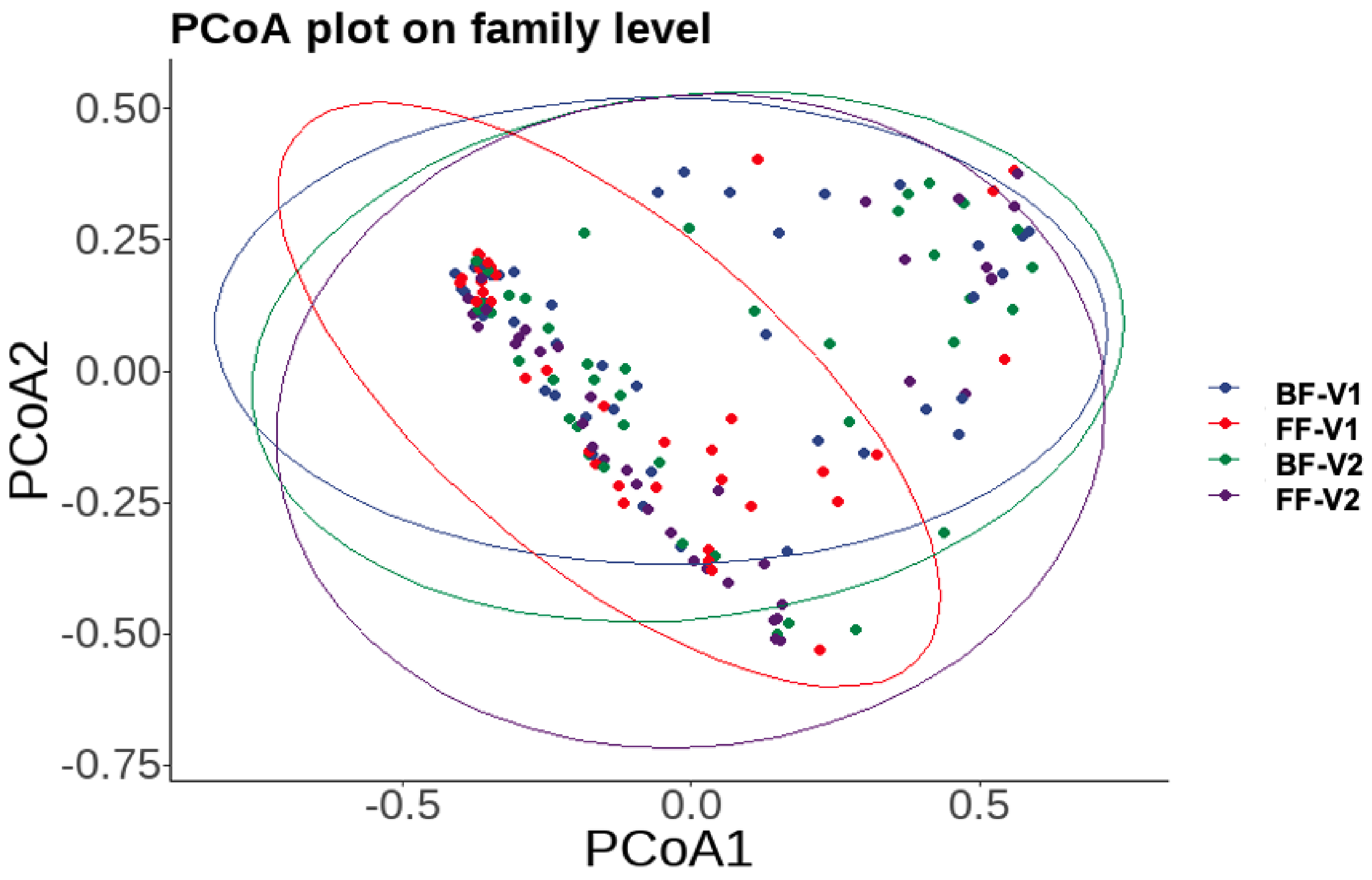


*Points of different colors represent samples of different groups.

**Additional file 8.** Relative proportion (%) of SCFAs and BCFAs for the full analysis set[1].

| Metabolite (% relative) | FF (n=60) | | BF (n=60) | | P-value |
|---|---|---|---|---|---|
| | V1 | V2 | V1 | V2 | |
| **Total BCFAs** | **0.5 (0.7)** | **1.1 (1.1)** | **0.6 (1.2)** | **0.7 (1.3)** | **0.048** |
| **Acetate** | **81.4 (14.4)** | **75.3 (12.1)** | **84.0 (12.5)** | **84.3 (12.4)** | **0.026** |
| **Propionate** | **9.5 (9.1)** | **13.7 (8.4)** | **7.8 (8.4)** | **8.4 (7.4)** | **0.012** |
| Butyrate | 9.2 (10.9) | 10.2 (10.7) | 9.0 (8.7) | 7.5 (9.6) | 0.526 |
| Valerate | 0.2 (0.3) | 0.2 (0.2) | 0.1 (0.2) | 0.2 (0.3) | 0.888 |
| Caproate | 0.04 (0.04) | 0.03 (0.02) | 0.08 (0.2) | 0.1 (0.3) | 0.246 |
| Isobutyrate | 0.2 (0.4) | 0.6 (0.6) | 0.3 (0.7) | 0.4 (1.0) | 0.299 |
| **Isovalerate** | **0.2 (0.4)** | **0.5 (0.5)** | **0.2 (0.5)** | **0.2 (0.4)** | **0.001** |

BCFA: Branched chain fatty acid; BF: Breastfed; FF: Formula-fed; SCFA: Short chain fatty acid; V1: Visit 1; V2: Visit 2.

[1]Data presented are mean (SD) % relative proportions. Analyzed (at V2) by ANCOVA models correcting for the corresponding site, baseline value, delivery method, sex, and baseline age. P-value results from a two-sided test of difference between the FF and BF groups at V2 estimated with ANCOVA.

**Additional file 9.** Fecal biomarkers of immune response, inflammation, and intestinal barrier integrity for the full analysis set.

| **Biomarker (Per dry feces)** | **FF (n=60)** | | **BF (n=60)** | | **P-value** |
|---|---|---|---|---|---|
| | **V1** | **V2** | **V1** | **V2** | |
| sIgA (mg/g) | 2.0 (1.6) | 2.4 (1.9) | 2.5 (2.8) | 3.4 (3.1) | 0.068 |
| IL-1β (ng/g) | 2.4 (0.8) | 3.2 (1.0) | 2.2 (0.5) | 4.7 (2.0) | 0.244 |
| IFN-γ (ng/g) | 43.1 (17.3) | 50.5 (21.6) | 37.7 (15.1) | 60.3 (33.5) | 0.072 |
| TNF-α (ng/g) | 2.7 (1.2) | 3.4 (1.3) | 2.4 (1.2) | 3.9 (2.3) | 0.164 |
| Calprotectin (μg/g) | 3.0 (2.1) | 6.0 (5.0) | 4.0 (4.1) | 4.4 (4.1) | 0.116 |
| α1antitrypsin (μg/g) | 842.9 (500.8) | 1221.2 (477.7) | 749.7 (443.0) | 1246.3 (679.2) | 0.601 |
| **Lipocalin-2 (μg/g)** | **21.7 (16.9)** | **29.0 (19.8)** | **16.3 (13.1)** | **36.3 (28.5)** | **0.048** |

BF: Breastfed; FF: Formula-fed; sIgA: Secretory immunoglobulin A; V1: Visit 1; V2: Visit 2.

[1]Data presented are mean (SD). Analyzed by robust ANCOVA models correcting for the corresponding site, baseline value, delivery method, sex, and baseline age. P-value results from a two-sided test of difference between the FF and BF groups at V2 estimated with robust ANCOVA.

**Additional file 10.** Spearman rank correlations between gut microbiota and fecal organic acids at visit 2 (V2) with Benjamini-Hochberg false discovery rate correction applied to the set of two-sided p-values.

| **Species** | **Acetic acid** | **Butyric acid** | **Propanoic acid** | **Total BCFAs** | **Total SCFAs** |
|---|---|---|---|---|---|
| Acinetobacter | -0.143 (FDR ρ=0.407) | -0.0124 (FDR ρ=0.92) | -0.185 (FDR ρ=0.345) | -0.303 (FDR ρ=0.083) | -0.154 (FDR ρ =0.407) |
| Actinomyces | 0.185 (FDR ρ=0.513) | -0.128 (FDR ρ=0.664) | 0.0289 (FDR ρ=0.958) | 0.00625 (FDR ρ=0.958) | 0.143 (FDR ρ=0.664) |
| Atopobium | 0.15 (FDR ρ=0.407) | -0.105 (FDR ρ=0.485) | -0.0542 (FDR ρ=0.646) | -0.0936 (FDR ρ=0.485) | 0.125 (FDR ρ=0.432) |
| Bacteroides | -0.0174 (FDR ρ=0.935) | -0.215 (FDR ρ=0.342) | 0.0222 (FDR ρ=0.935) | 0.0332 (FDR ρ=0.935) | -0.0268 (FDR ρ=0.935) |
| Bifidobacterium | **0.496 (FDR ρ<0.0001)** | -0.203 (FDR ρ=0.143) | 0.0891 (FDR ρ=0.506) | 0.216 (FDR ρ=0.119) | **0.407 (FDR ρ=0.001)** |
| Blautia | 0.267 (FDR ρ=0.115) | -0.103 (FDR ρ=0.514) | 0.168 (FDR ρ=0.342) | 0.126 (FDR ρ=0.514) | 0.25 (FDR ρ=0.115) |
| Citrobacter | 0.208 (FDR ρ=0.169) | 0.262 (FDR ρ=0.129) | 0.236 (FDR ρ=0.129) | 0.135 (FDR ρ=0.386) | 0.27 (FDR ρ=0.129) |
| Clostridium | **-0.319 (FDR ρ=0.017)** | **0.498 (FDR ρ<0.001)** | -0.0674 (FDR ρ=0.639) | -0.178 (FDR ρ=0.198) | -0.24 (FDR ρ=0.071) |
| Collinsella | 0.103 (FDR ρ=0.429) | -0.211 (FDR ρ=0.306) | 0.125 (FDR ρ=0.429) | 0.155 (FDR ρ=0.342) | 0.109 (FDR ρ=0.429) |
| Cutibacterium | 0.134 (FDR ρ=0.882) | 0.104 (FDR ρ=0.882) | 0.0486 (FDR ρ=0.882) | 0.0503 (FDR ρ=0.882) | 0.12 (FDR ρ=0.882) |
| Dysgonomonas | 0.187 (FDR ρ=0.141) | 0.293 (FDR ρ=0.065) | 0.31 (FDR ρ=0.065) | 0.227 (FDR ρ=0.087) | 0.247 (FDR ρ=0.087) |
| Eggerthella | 0.127 (FDR ρ=0.637) | -0.134 (FDR ρ=0.637) | 0.021 (FDR ρ=0.966) | 0.0677 (FDR ρ=0.732) | 0.079 (FDR ρ=0.732) |
| Enterobacter | -0.108 (FDR ρ=0.627) | 0.139 (FDR ρ=0.576) | -0.00788 (FDR ρ=0.947) | -0.153 (FDR ρ=0.576) | -0.0819 (FDR ρ=0.627) |
| Enterococcus | 0.126 (FDR ρ=0.366) | 0.137 (FDR ρ=0.366) | 0.21 (FDR ρ=0.217) | 0.24 (FDR ρ=0.193) | 0.167 (FDR ρ=0.349) |
| Erysipelatoclostridium | 0.0251 (FDR ρ=0.936) | -0.129 (FDR ρ=0.438) | -0.0846 (FDR ρ=0.609) | 0.131 (FDR ρ=0.438) | -0.000543 (FDR ρ=0.996) |

| | | | | | |
|---|---|---|---|---|---|
| Escherichia | 0.216 (FDR ρ=0.430) | 0.0895 (FDR ρ=0.839) | 0.0545 (FDR ρ=0.957) | 0.0889 (FDR ρ=0.839) | 0.195 (FDR ρ=0.430) |
| Finegoldia | -0.0683 (FDR ρ=0.908) | -0.0111 (FDR ρ=0.983) | -0.202 (FDR ρ=0.378) | -0.0468 (FDR ρ=0.908) | -0.0991 (FDR ρ=0.902) |
| Flavonifractor | -0.137 (FDR ρ=0.818) | -0.054 (FDR ρ=0.818) | 0.0388 (FDR ρ=0.818) | 0.0787 (FDR ρ=0.818) | -0.107 (FDR ρ=0.818) |
| Fusicatenibacter | 0.261 (FDR ρ=0.161) | -0.185 (FDR ρ=0.290) | 0.0724 (FDR ρ=0.613) | 0.0871 (FDR ρ=0.613) | 0.225 (FDR ρ=0.161) |
| Gemella | 0.0809 (FDR ρ=0.555) | -0.158 (FDR ρ=0.249) | **-0.357 (FDR ρ=0.007)** | **-0.286 (FDR ρ=0.032)** | -0.00832 (FDR ρ=0.944) |
| Gordonibacter | 0.127 (FDR ρ=0.634) | -0.141 (FDR ρ=0.634) | 0.048 (FDR ρ=0.789) | 0.0457 (FDR ρ=0.789) | 0.0767 (FDR ρ=0.789) |
| Haemophilus | -0.0303 (FDR ρ=0.798) | -0.0848 (FDR ρ=0.627) | -0.244 (FDR ρ=0.066) | **-0.267 (FDR ρ=0.050)** | -0.0955 (FDR ρ=0.627) |
| Hungatella | 0.000508 (FDR ρ=0.997) | -0.123 (FDR ρ=0.997) | 0.00821 (FDR ρ=0.997) | -0.0112 (FDR ρ=0.997) | -0.0133 (FDR ρ=0.997) |
| Klebsiella | -0.146 (FDR ρ=0.387) | **0.344 (FDR ρ=0.017)** | 0.109 (FDR ρ=0.529) | -0.0427 (FDR ρ=0.810) | -0.0663 (FDR ρ=0.738) |
| Lachnoclostridium | 0.00969 (FDR ρ=0.958) | -0.0065 (FDR ρ=0.958) | 0.114 (FDR ρ=0.601) | 0.177 (FDR ρ=0.402) | 0.0297 (FDR ρ=0.958) |
| Lactobacillus | 0.269 (FDR ρ=0.170) | 0.0544 (FDR ρ=0.817) | 0.169 (FDR ρ=0.453) | 0.106 (FDR ρ=0.817) | 0.242 (FDR ρ=0.170) |
| Lactococcus | 0.232 (FDR ρ=0.210) | 0.109 (FDR ρ=0.558) | 0.189 (FDR ρ=0.238) | 0.191 (FDR ρ=0.238) | 0.237 (FDR ρ=0.210 |
| Megamonas | 0.138 (FDR ρ=0.320) | -0.226 (FDR ρ=0.223) | 0.209 (FDR ρ=0.223) | 0.137 (FDR ρ=0.320) | 0.155 (FDR ρ=0.320) |
| Parabacteroides | -0.0398 (FDR ρ=0.828) | -0.227 (FDR ρ=0.243) | 0.102 (FDR ρ=0.584) | 0.106 (FDR ρ=0.584) | -0.0252 (FDR ρ=0.831) |
| Prevotella | 0.0585 (FDR ρ=0.699) | -0.0623 (FDR ρ=0.699) | -0.0889 (FDR ρ=0.699) | 0.0406 (FDR ρ=0.733) | 0.0609 (FDR ρ=0.699) |
| Raoultella | 0.0868 (FDR ρ=0.756) | 0.112 (FDR ρ=0.756) | 0.0897 (FDR ρ=0.756) | 0.0858 (FDR ρ=0.756) | 0.103 (FDR ρ=0.756) |
| Rothia | -0.211 (FDR ρ=0.079) | **-0.383 (FDR ρ=0.002)** | **-0.602 (FDR ρ<0.0001)** | **-0.514 (FDR ρ<0.0001)** | **-0.326 (FDR ρ=0.006)** |
| Ruthenibacterium | 0.0518 (FDR ρ=0.756) | -0.186 (FDR ρ=0.284) | 0.0641 (FDR ρ=0.850) | 0.186 (FDR ρ=0.284) | 0.0353 (FDR ρ=0.861) |

| | | | | | |
|---|---|---|---|---|---|
| Serratia | 0.0685 (FDR ρ=0.632) | -0.177 (FDR ρ=0.290) | -0.074 (FDR ρ=0.632) | -0.179 (FDR ρ=0.290) | 0.0247 (FDR ρ=0.835) |
| Staphylococcus | -0.109 (FDR ρ=0.455) | -0.0393 (FDR ρ=0.842) | **-0.305 (FDR ρ=0.026)** | **-0.316 (FDR ρ=0.026)** | -0.153 (FDR ρ=0.291) |
| Streptococcus | 0.1 (FDR ρ=0.532) | 0.1 (FDR ρ=0.532) | **-0.272 (FDR ρ=0.034)** | **-0.295 (FDR ρ=0.034)** | 0.0501 (FDR ρ=0.671 |
| Veillonella | 0.121 (FDR ρ=0.456) | 0.162 (FDR ρ=0.333) | 0.255 (FDR ρ=0.196) | 0.0539 (FDR ρ=0.732) | 0.167 (FDR ρ=0.333) |
